# Learning phenotypic patterns in genetic diseases by symptom interaction modeling

**DOI:** 10.1101/2022.07.29.22278181

**Authors:** Kevin Yauy, Nicolas Duforet-Frebourg, Quentin Testard, Sacha Beaumeunier, Jerome Audoux, Benoit Simard, Dimitri Larue, Michael G. B. Blum, Virginie Bernard, David Genevieve, Denis Bertrand, PhenoGenius consortium, Nicolas Philippe, Julien Thevenon

## Abstract

Observing phenotyping practices from an international cohort of 1,686 cases revealed heterogeneity of phenotype reporting among clinicians. Heterogeneity limited their exploitation for diagnosis as only 43% of symptom-gene associations in the cohort were available in public databases. We developed a symptom interaction model that summarized 16,600 terms into 390 groups of interacting symptoms and detected 3,222,053 novel symptom-gene associations. By learning phenotypic patterns in genetic diseases, symptom interaction modeling handled heterogeneity in phenotyping, to the extent of covering 98% of our cohort’s symptom-gene associations. Using these symptom interactions improved the diagnostic performance in gene prioritization by 42% (median rank 80 to 41) compared to the best algorithms. Symptom interaction modeling will provide new discoveries in precision medicine by standardizing clinical descriptions.

**One sentence summary:** Learning phenotypic patterns in genetic disease by symptom interaction modeling addresses physicians’ heterogeneous phenotype reporting.

---

Precision medicine relies on patient stratification and recognition of clinically relevant groups to improve diagnosis, prognosis, and medical treatment ^1^. Phenotyping allows homogeneous groups of individuals to be constituted, where physicians report characteristics deviating from normal morphology, physiology, and behavior using standardized descriptions in the Human Phenotype Ontology (HPO) ^2,3^. Despite a common ontology and abundant clinical data, medical records often lack consistency and comparability between descriptions and practitioners, which is referred to as fuzzy matching in phenotype profiles ^4^. This inconsistent phenotyping is a major hurdle to fully exploiting the clinical data contained in medical records. Nevertheless, no studies about phenotyping practices in clinical sequencing are known to have been undertaken until now.

## 1. Phenotyping practices in large cohorts

Through four international studies, including 1,686 patients in total, we collected 2501 different symptoms in HPO format and 849 different disease-causing genes ^5–7^ (Table S1). Nearly half of the patients in the multi-center cohort had symptoms belonging to the *Abnormality of the nervous system* (HP:0000707) and *Abnormality of the musculoskeletal system* (HP:0033127) classes, illustrating the current focus on those rare disorders in clinical practice ^8^ (Figure S1). Reflecting the genetic heterogeneity of rare diseases, 538 of 849 genes were declared only once in the cohort and the most frequently mutated gene occurred in less than 2% of cases (*ABCC6*,n=21, Table S2).

We observed heterogeneity in HPO selection terms, as 47% of terms were used only once (Figure 1A, Table S3). The median number of HPO terms per physicians’ clinical description varied across observations, ranging from three (Peng *et al*. ^7^) to seven (PhenoGenius consortium, Seo *et al*. and Trujillano *et al*. ^5,6^) (Figure 1B). The heterogeneity of physicians’ clinical descriptions was also observed for patients with identical genetic diagnoses. For genes involved in diagnosis of more than ten patients, 67 % of symptoms were declared in only one clinical description.

**Fig 1.**
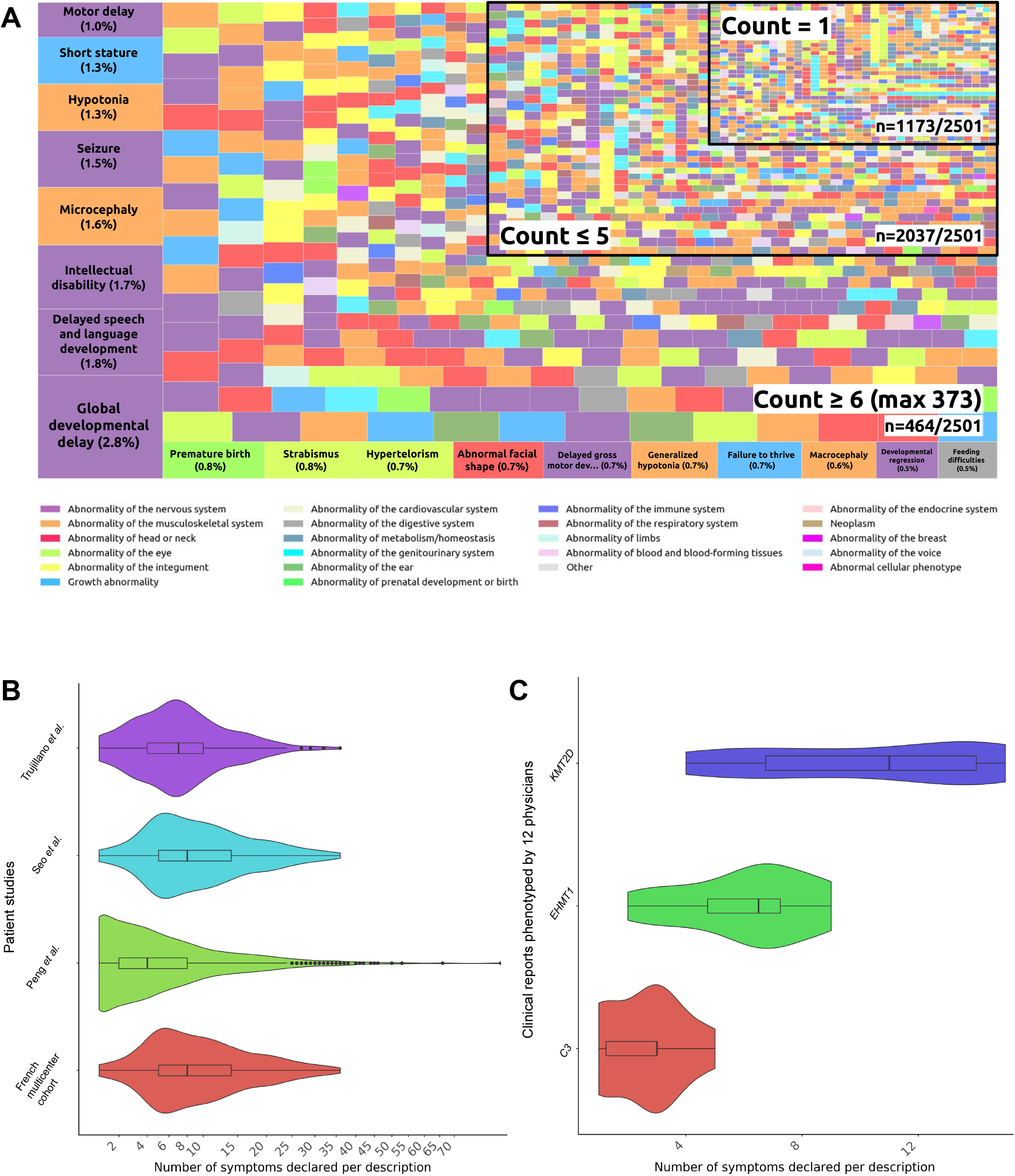
The landscape of phenotyping practices from a retrospective cohort of 1,686 patients and a prospective experiment of clinical reports phenotyped by multiple physicians. A. Treemap chart of the HPO terms frequency across the retrospective cohort. B. Violin plot of HPO term counts per clinical description for each subgroup of the cohort. C. Violin plot of HPO term counts per clinical description for each clinical report phenotyped by 12 physicians.

To exclude the possibility that the observed heterogeneity was due to variability in clinical examinations, we next investigated whether heterogeneity in clinical descriptions was reported if physicians phenotyped the same clinical observations. We settled on a prospective experiment where 12 clinical geneticists with various levels of expertise (Table S4) were asked to phenotype three independent clinical reports associated with genetic test prescription, i.e. to convert free text to phenotypes in HPO format. We observed heterogeneity in terms of the number and diversity of symptoms declared per clinical observation (Figure 1C). For instance, two to nine symptoms were declared in clinical descriptions of the Kleefstra syndrome observation with the *EHMT1* pathogenic variant. A total of 29 different terms were provided; 17 of these terms were used by two or more physicians, and none of the terms were mentioned by all 12 physicians.

## 2. Quantifying the overlap of symptoms-gene associations between the retrospective cohort and the medical literature

To assess if the clinical descriptions of our cohort matched available knowledge in the medical literature, we mapped the cohort’s 11,526 unique symptom-gene associations to the 734,931 associations available in HPO-structured databases (Orphanet, DDG2P ^9^, and the Monarch Initiative or MI ^3^). From these databases, only 4,913 associations (43%) matched, meaning that 57% were missing (Figure 2A).

**Fig 2.**
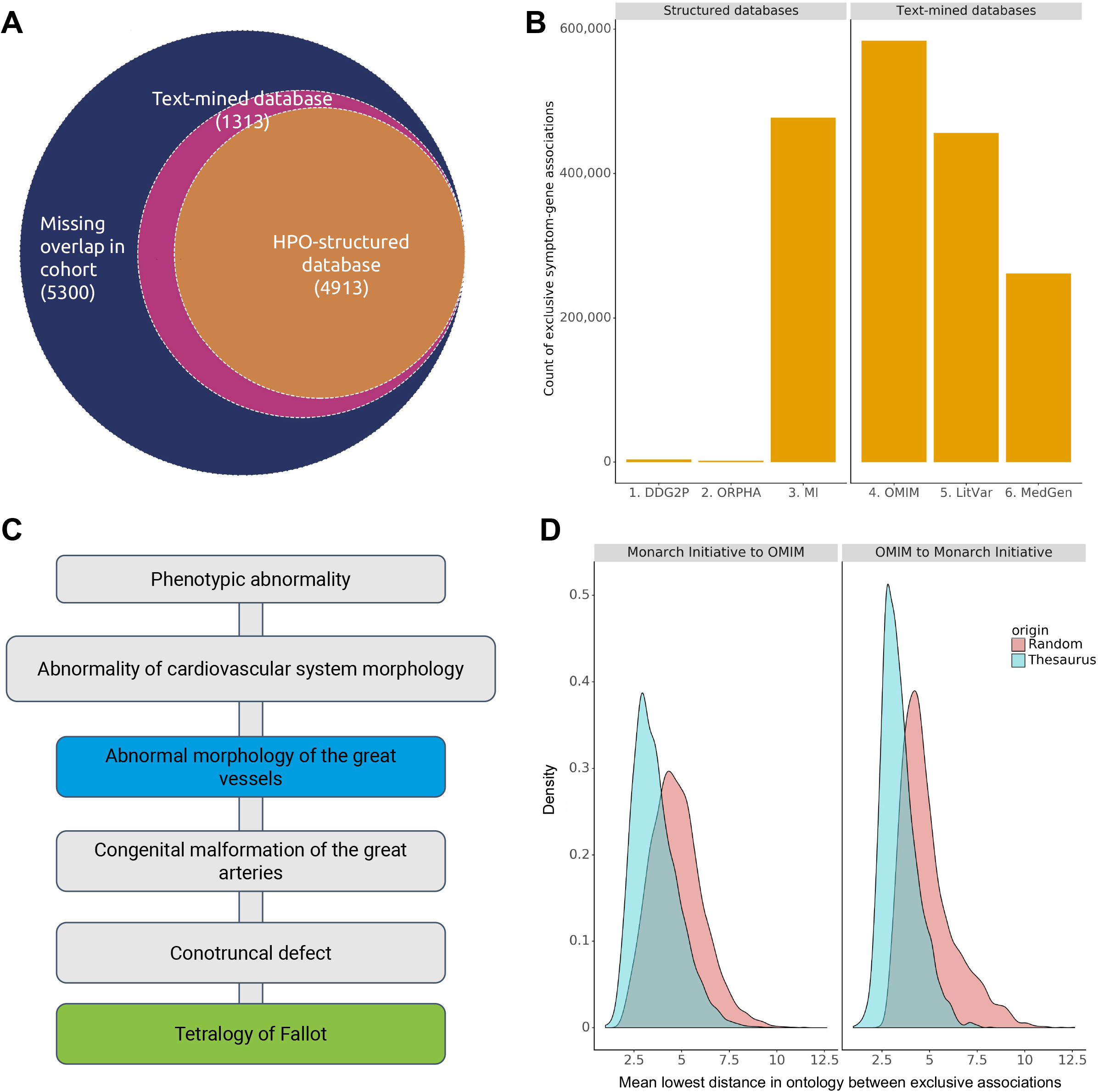
Quantifying the overlap of symptoms-gene associations between the retrospective multicenter cohort of 1,686 patients and the medical literature. A. Venn diagram of symptom-gene associations observed in cohort overlapped with public HPO-structured databases and text-mined associations in free-text databases. B. Count distribution of symptoms-gene association exclusive to each database. C. Illustration of exclusive symptom-gene associations found in Monarch Initiative database (blue) and text-mined OMIM database (green), using *KMT2D* as an example. Gray associations were unfound. D. Distribution of the mean lowest distance in the ontology between exclusive terms in the Monarch Initiative database and our text-mined OMIM database, compared to a random choice of HPO terms.

As the clinical descriptions of genetic diseases in medical literature are mainly available in free-text format, we developed a text-mining algorithm based on Elasticsearch to extract symptom-gene associations from free-text data in HPO format. Applied to OMIM ^10^, MedGen ^11^, and abstracts from PubMed, this text-mining algorithm identified an additional 1,049,522 symptom-gene associations. This approach resulted in a 3.2-fold increase in HPO-structured database associations (Figure 2B).

The text-mining algorithm provided symptom-gene associations where symptoms were significantly deeper in the ontology compared to the HPO-structured databases (median depth 6.7 and 5.2 respectively, Kolmogorov-Smirnov test p-value < 10^−215^, Figure S2). This underlines the complementarity of these approaches, as illustrated in Figure 2C where *KMT2D* was associated with *Abnormal morphology of the great vessels* (HP:0030962) in the MI database and *Tetralogy of Fallot* (HP:0001636) in the OMIM database. Reflecting the variability across individuals in selecting an HPO term to summarize a clinical observation, 76% of associations were exclusive to one database. We hypothesized that text-mined symptom-gene associations in the literature were related to associations available in HPO-structured databases. This hypothesis embodies the fuzzy phenotyping concept, providing human-determined alternative wordings of the same information.

To evaluate this hypothesis, for each gene we compared the average distance in the ontology of exclusive symptom-gene associations to the MI database and the text-mined OMIM database, respectively the largest database of each type (Figure 2B). Compared with a random choice of an HPO term, the average distance of the exclusive symptom-gene associations was significantly lower, suggesting these associations are related (Kolmogorov-Smirnov test p-value < 10^−215^, Figure 2D).

Although in this exercise the number of symptom-gene associations increased from 734,931 (MI, DDG2P, Orphanet database) to 1,784,453 (with associations found with the text mining algorithm), a match with the cohort’s symptom-gene associations was only available for 6,226 of 11,526 (57%) associations, meaning that 43% of matches were still missing (Figure 2A).

## 3. From symptom-gene to symptom-symptom associations modeling

We investigated whether modeling associations between symptoms of the same genetic disorder improved matches. As the Human Phenotype Ontology is ordered according to human development, it may not represent the interaction of symptoms in disease (Figure 3A). We explored an alternative approach to measure symptom-symptom associations in genetic diseases. We considered a node similarity algorithm based on a knowledge graph that stored the symptom-gene associations we collected from the literature.

**Fig 3.**
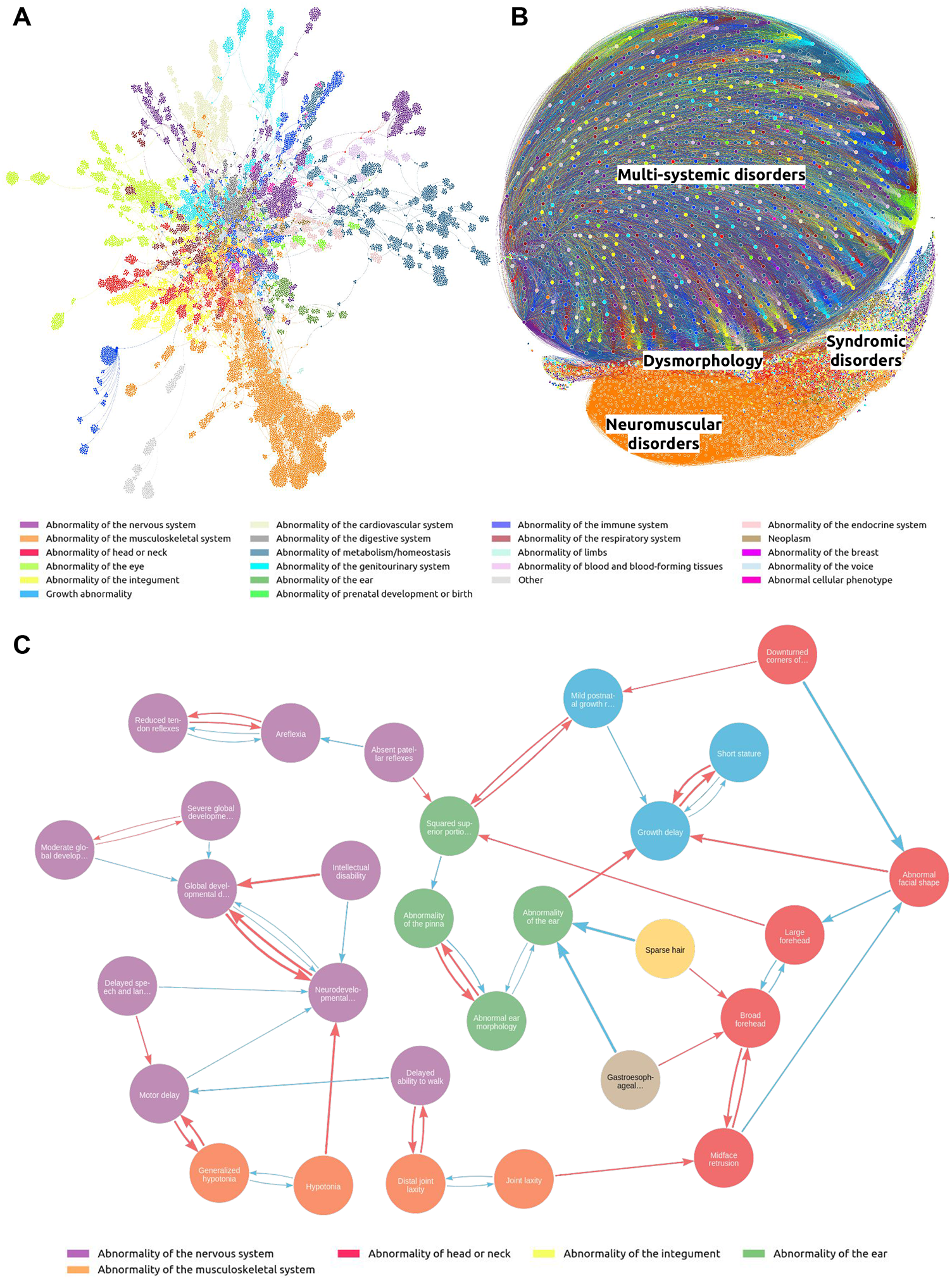
Modeling symptom-symptom interaction in rare diseases using node similarity algorithms on collected symptoms-gene associations. Node color represents the main HPO class. A. Graph visualization of symptom relationships based on the human development architecture of HPO. B. Graph visualization of symptoms relationships with node similarity > 80%. C. Illustration of symptom relationships with the Kleefstra syndrome clinical report with *EHMT1* variant, phenotyped by 12 geneticists. Blue arrows linked the closest symptom in HP ontology and red arrows the symptom with the highest node similarity among declared symptoms.

We found a high correlation between symptom-symptom similarity pair scores and their frequency of co-occurrence in clinical observations (Spearman correlation coefficient: 0.99). No correlation was observed between symptom-symptom similarity pair scores and the distance between symptoms in the HPO (Spearman correlation coefficient: −0,02, Figure S3), reflecting that symptom-symptom associations cannot be solely derived from the ontology architecture.

According to similarity score distributions, we posited that similarities above 80% were potential substitutes or highly similar symptoms in diseases (Figure S4). This resulted in the selection of 565,943 pairs of highly similar symptoms, corresponding to the 10% highest symptom-symptom association scores (Figure 3B). A total of 26% of these pairs were observed for symptoms in the same ontology class (145,611 of 565,943), mostly from the *Abnormality of the musculoskeletal system* (HP:0033127) class (51%, 73,817 of 145,611). Inter-classes pairs of symptoms represented 74% of highly similar symptoms, where the most recurrent pair was *Abnormality of metabolism/homeostasis* (HP:0001939) with *Abnormality of the nervous system* (HP:0000707) (8%, 35,476 of 420,332).

We illustrate these similarities in Figure 3C, using the symptom *Hypotonia* (HP:0001290) reported by six of the 12 practitioners in our exercise on the Kleefstra syndrome with the *EHMT1* pathogenic variant. In the symptom-symptom association graph, the closest term to *Hypotonia* is *Neurodevelopmental delay* (HP:0012758), with a symptom-symptom similarity pair score measuring 86%. In the HPO, these symptoms are separated by ten nodes and belong to two different main classes: *Abnormality of the musculoskeletal system* (HP:0033127) and *Abnormality of the nervous system* (HP:0000707) respectively.

We then investigated to what extent considering two highly similar symptoms as substitutes improved the coverage of symptoms-gene associations. Among the cohort’s 11,526 unique symptom-gene associations, only 6,226 associations were found in HPO-structured and text-mined databases, but this number rises to 8,350 when accounting for similarities. Considering substitutes provided additional 1,506,469 symptom-gene associations to the previous 1,784,453 associations from MI, DDG2P, Orphanet, and text-mined databases.

Modeling associations between symptoms revealed a majority of inter-HPO classes included similar symptoms, highlighting the missing aspect of symptom relationships in the HP ontology. Enhancing symptoms with their highly similar pairs improved coverage of symptom-gene associations in the cohort, but 27% of associations were still missing.

### 4. From symptom-gene associations to groups of symptoms modeling

Symptom-symptom associations were evaluated independently when identifying substitutes based on node similarity. To gain better coverage of symptom-gene associations, we considered a more elaborate collaborative filtering approach based on non-negative matrix factorization (NMF) ^12^.

Using the topic coherence measure ^13^, we determined that the 16,660 HPO terms could optimally be reduced to 390 groups of interacting symptoms or phenotypic patterns (Figure S5). Each symptom was positioned in the graph with group weights determined by the algorithm (Figures 4A-4B). Each gene was associated in a median of 36 groups and a group with a median of 501 genes. To compare the recall of the NMF and the node similarity model, we kept only the top 10% of 390 symptom-groups weights (Figure S6). Overall in this selection, there were 43,308 symptom-group associations leading to 5,971,755 pairs of symptoms.

**Fig 4.**
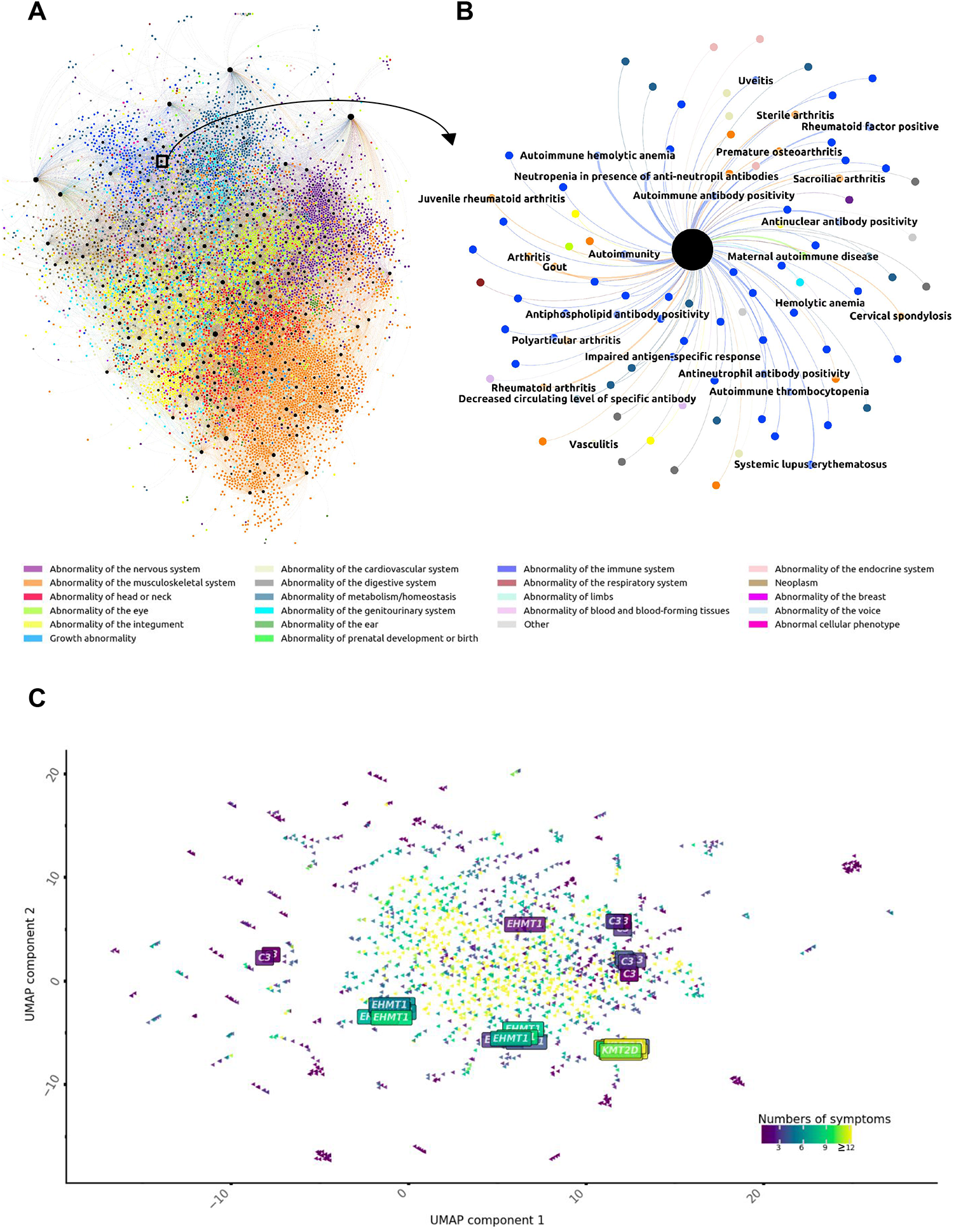
Modeling symptom-symptom interactions in genetic disease using non-negative matrix factorization. A. Visualization of symptom relationship based on 390 groups of interacting symptoms from medical literature. Group 273 is highlighted by the black box and arrow. B. Illustration of group 273 with the main symptom *Autoimmunity* (HP:0002960). For graphs in figures A and B, the line thickness is proportional to the weights of symptoms in the group. Colors correspond to the main HPO class and groups are in black. For readability, only the top 10% of symptom-group associations are displayed. C. UMAP visualization of cohort’s clinical descriptions projected into the group of symptom dimension, colored by the number of symptoms. Boxes represent clinical reports description phenotyped by twelve physicians.

We investigated to what extent the coverage of symptoms-gene associations was improved by considering that two symptoms belonging to the same group were substitutes. Using these pairs of symptom associations enhanced the coverage of symptom-gene associations to 11,340 of the 11,526 associations from the cohort, leaving less than 2% of matches missing. This new manner of detecting associations resulted in the addition of 2,163,663 NMF-based symptom-gene associations to the previous 1,784,453 associations obtained from MI, DD2P, Orphanet, and text-mined databases. NMF-based symptom-gene associations overlapped with 99% of similarity-based associations (1,497,601 of 1,506,469).

To evaluate if these 390 phenotypic patterns represented the clinical spectrum of genetic diseases, we projected the cohort into the groups of symptoms dimension and performed a UMAP visualization ^14^. We applied agglomerative clustering to the cohort and compared clustering patient performance using this projection and the 16,600 HPO dimension. Using the initial list of 16,600 symptoms, 152 patients were found in 14 clusters significantly enriched in symptoms (Fisher exact test with p-value < 0.05 with Benjamini Hochberg correction) (Figures S7-S8). Applying the projection in groups of symptoms, 1,136 patients were found in 51 clusters significantly enriched in groups of symptoms (Figure S9, Figure S10). To evaluate if this projection could standardize clinical descriptions, we applied it to the three clinical reports phenotyped by the 12 physicians in our experiment. We demonstrated the high coherence of our method even with symptom heterogeneity when sufficient numbers of HPO terms were given (*KMT2D report*) (Figure 4C, Figure S11). When fewer than 5 terms were provided, clinical description projections still grouped patients but with lower homogeneity (*EHMT1,C3*).

The delineation of 390-groups of interacting symptoms enabled an increase in coverage of the available knowledge on genetic disorders and provided a way of building on HP ontology to standardize clinical descriptions. Next, we used symptom interaction modeling to develop a phenotype matching system.

### 5. Symptom interaction models as an efficient and robust system for phenotype matching

To evaluate the clinical relevance of symptom interaction models, we designed phenotype matching and diagnostic gene ranking experiments. We defined a phenotype match when at least one symptom in the clinical description was related to the diagnostic gene (Figure 5A). According to the count of matches per gene, a personalized ranked list of genes was provided (Figure S12). These experiments were performed on the clinical observations of 1,686 patients.

**Fig 5.**
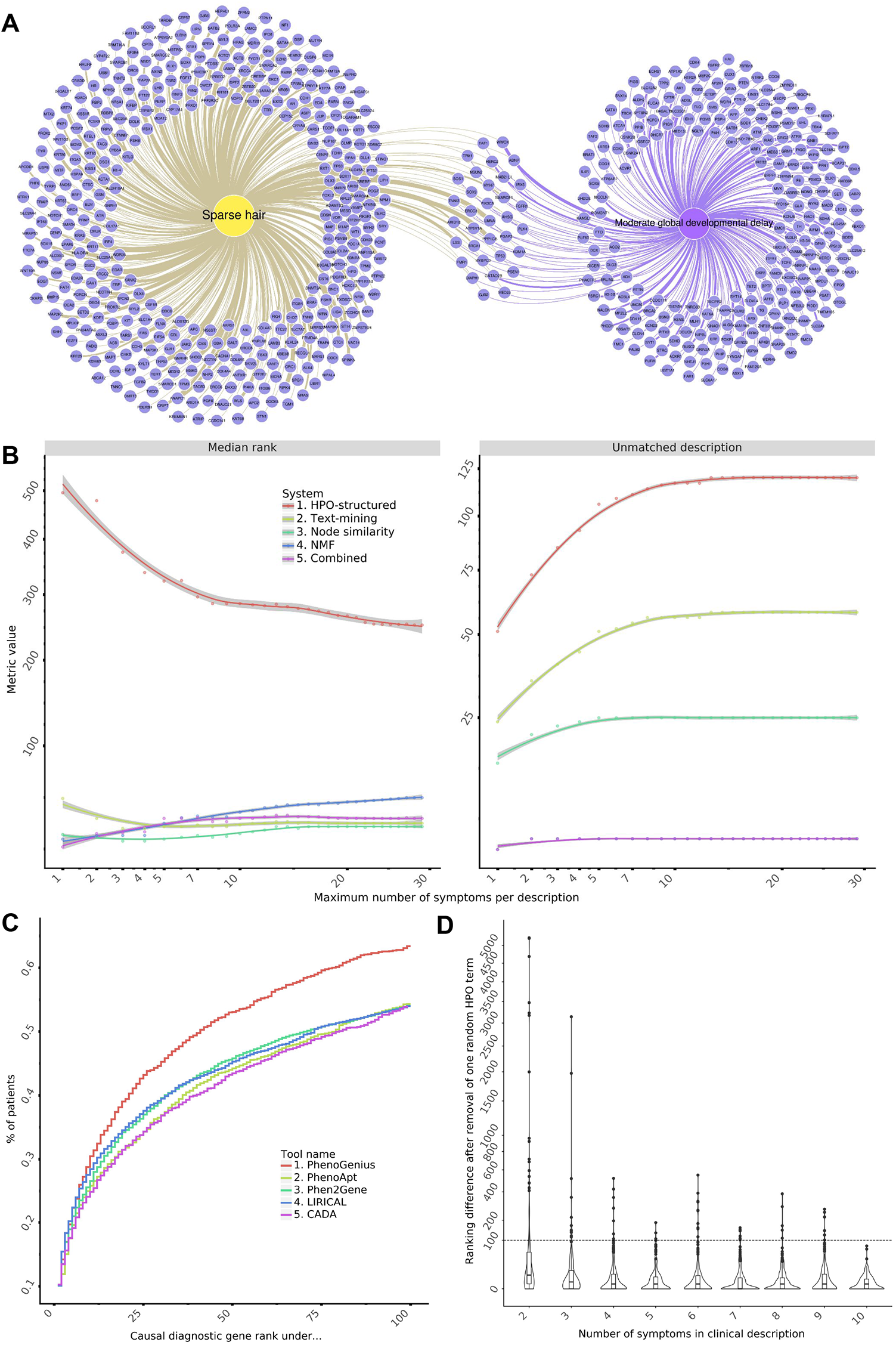
Modeling symptom interactions as an efficient system for phenotype matching. A. Illustration of the principle of phenotype matching, looking for the most connected genes to the clinical description containing two symptoms of the Kleefstra syndrome observation with the *EHMT1* variant: *Sparse hair* (HP:0008070, yellow) and *Moderate global developmental delay* (HP:0011343, purple). Line thickness is proportional to the probability score of symptom-gene associations available with joint HPO-structured and text-mined databases. B. Performance benchmark metrics of diagnostic gene prioritization ranking (median rank, left side) and phenotype matching (count of unmatched description, right side) according to a maximum number of symptoms in clinical descriptions of the cohort. C. Benchmark of a selection of state-of-the-art gene prioritization programs. The fraction of cases correctly diagnosed (y-axis) is plotted against a cumulative causal gene rank. D. Ranking differences after removing one symptom according to the number of terms in clinical descriptions.

Using the HPO-structured databases (MI, DDG2P, Orphanet), we obtained a phenotype match for 1,566 clinical observations with a median diagnostic gene rank of 251. Applying text-mined associations led to a match for 1,628 clinical observations with a median rank of 40 (Figure 5B). The best performance in median diagnostic rank was provided by node similarity symptoms association (median rank 37, compared to 58 with NMF), but NMF was able to get a more exhaustive coverage of clinical observations (1682, compared to 1663 with node similarity). This coverage gap was exclusively observed where the clinical descriptions contained five terms or less (four unmatched descriptions, compared to 25 with node similarity). As each symptom interaction model provides a different level of inductive reasoning, we conditionally applied a model according to the number of symptoms in the clinical description. The combined system, which we called PhenoGenius provided the best performance (median rank 41) and reached a nearly full phenotype match of diagnostic genes (99.8%, 1682/1686) for all clinical descriptions.

To illustrate this phenotype-matching system, we considered a clinical description containing two symptoms of the Kleefstra syndrome observation with the *EHMT1* pathogenic variant: *Sparse hair* (HP:0008070) and *Moderate global developmental delay* (HP:0011343). There is no match between these terms and *EHMT1* in HPO-structured databases. No match is identified from text-mined symptom-gene associations either. Symptom interaction modeling achieves phenotype matches, ranking 1244 out of 5235 (top 25% of genes) with the similarity model and 851 out of 5235 (top 17% of genes) with the NMF model and PhenoGenius combined system.

We then compared PhenoGenius to four recently published algorithms for phenotype-driven gene prioritization: PhenoApt, Phen2Gene, CADA, and LIRICAL ^7,15–17^. Despite using different prioritization methodologies, these four programs demonstrated similar performances in phenotype matching (Figure 5C). Using symptom interaction modeling, PhenoGenius (median rank 41) increased the median diagnostic gene rank by 42% compared to the best competitor, Phen2Gene (median rank 71, 73 to 80 for other methods). This improvement was replicated across each study subgroup in the cohort, highlighting the clinical relevance of symptom interactions in genetic disease models (Figure S13).

To assess the robustness of gene prioritization, we randomly removed each symptom from clinical descriptions with two terms or more and measured the consequence on the disease-causing gene ranking for descriptions in the top-ranked half of the cohort (rank 41 or lower). Overall, 701 clinical descriptions led to 6,331 symptom removal experiments. In most cases, phenotype matching remained robust with symptom removal (Figure 5D). Disease-causing gene ranking was identical in 35% of cases (2,274 of 6,331) and the median of absolute differences between ranks was only one. However, nine extreme drops in the ranking (> 1000) were observed with clinical descriptions with three or fewer terms, including two complete loss of phenotype matches for descriptions with two symptoms. For clinical descriptions with four or more terms, we found no extreme drops in gene rankings.

## Discussion

This study used symptom interaction modeling to learn phenotyping patterns in genetic diseases. This method adds to the precision medicine toolbox with a way of standardizing clinical descriptions and matching physicians’ phenotyping to the medical knowledge of genetic diseases.

This study provides an in-depth analysis of phenotyping clinical practice by analyzing 1,686 phenotyping reports of patients with a definitive genetic diagnosis ^5–7^. In addition, a qualitative comparison of three clinical reports phenotyped by 12 physicians was performed. Complementary to recent reports ^8,18^, this study provides original insights on heterogeneous patient phenotyping, both in the cohort’s clinical descriptions and the medical literature. In our qualitative experiment, the main observation was the diversity of terms chosen by physicians to describe the exact same clinical description. These observations suggest that clinical description should be standardized, following harmonization of symptom description with HP ontology.

As well as encouraging richness of clinical description, tools must address the medical reality of summarized or partial clinical information. Lacking time or omitting symptoms in their clinical routine, physicians provide scanty phenotyping. Symptoms may be chosen based on strong clinical *a priori* or learned phenotypic patterns. Medical inductive knowledge often proposes patterns or groups of hypotheses based on recurrently associated symptoms in the physician’s own experience and in the literature. Defining groups of symptoms represent a natural behavior of medical inductive reasoning ^19,20^. This could explain the heterogeneity of phenotyping across clinical observations, independently from the innate clinical heterogeneity of a disorder.

To handle heterogeneous phenotyping, we developed symptom interaction models to standardize clinical descriptions and evaluate their clinical relevance through gene prioritization experiments. Based on symptom interaction models, PhenoGenius decreases the rank of the diagnostic gene by 42% compared to the best competitor. Its simplicity in scoring allows a complete understanding of phenotype matching, thus providing an interpretable measure of potential genotype-phenotype correlation. To lower the risk of missing a phenotype match because of a fuzzy description, clinical descriptions with four or more terms are recommended. Our approach contrasts with state-of-the-art phenotype-driven gene prioritization software, which mostly relies on complex scoring or symptom relationships based on HPO architecture.

Current algorithms address phenotyping heterogeneity using the ontology structure either to extract additional symptom-gene associations from literature or to evaluate the semantic similarity of symptoms ^21^. In contrast to these approaches, we used HPO as a dictionary of symptoms and considered relationships between symptoms only through their co-occurrence in genetic diseases found in HPO-structured and text-mined databases. Our algorithm uncovered the missing pieces of medical inductive reasoning in clinical descriptions through symptom similarity modeling and collaborative filtering using NMF methods ^12^. As such, projection into the symptoms interaction model dimension could provide a path to standardizing clinical descriptions. Moreover, the application of this algorithm is reproducible and interpretable, and these features are fundamental in a medical context ^22^. In addition, node similarity and NMF allow free association of symptoms, which is important since the same symptom may belong to different disease groups.

Our AI system performed well for gene prioritization. However, evaluation of our system’s performance in detecting gene/symptom associations is incomplete. In our international cohort, only 43% of symptom-gene associations were described in public databases. We have shown that the recall rate (percentage of detected associations among known associations) increased when considering similarity measures or techniques based on NMF. However, as the list of associations increased, an increase in recall came at a price of reduced precision, i.e. a reduced proportion of true associations among the detected associations. Evaluation of precision is impossible because some true associations are missing, highlighting the need to improve data sharing of physicians’ phenotype information.

As current knowledge overwhelms human learning abilities, an overarching goal in precision medicine is to overcome digital bottlenecks to succeed in deep phenotyping and identification of clinically relevant groups of patients. Progressive adoption of the Monarch Initiative’s HPO in clinical symptoms description, the development of automatic extraction of symptoms in HPO format from electronic medical records ^23^, and the definition of the Phenopackets standard file format by GAG4H ^24^ bring the community one step forward. A current challenge is integrating multiple data sources from electronic health records for deep phenotyping ^25^. Complementary to this challenge, we seek to standardize and improve the exploitation of clinical descriptions available in clinical practices using symptom interaction models. Long-standing aspirations are to be able to answer the question, “Have I seen a case like that before?” among extensive clinical data, and to identify undescribed symptom-gene associations ^26^.

Clinical description standardization using symptom interaction modeling may overcome several clinical bottlenecks in precision medicine. PhenoGenius is open-source, accessible through an interactive graph browser (https://github.com/kyauy/PhenoGenius), and a web app (https://phenogenius.streamlitapp.com/). This work paves the way for a set of tools to help identify new genes in disease, expand their clinical spectrum, and provide an easily interpretable clinical decision support system. If we can successfully deal with fuzzy phenotypic profiles and inductive medical reasoning in rare diseases, clinical data can be used for computational phenotype analysis, to improve the feasibility of precision medicine, and to support the adoption of genomic medicine.

## Data Availability

All data produced are available in Supplementary Materials and online at https://github.com/kyauy/PhenoGenius

## Data availability

The PhenoGenius source code is available for resource generation and scientific experiments in Apache License 2.0, including an interactive graph browser, on GitHub (https://github.com/kyauy/PhenoGenius). A web app is accessible at https://phenogenius.streamlitapp.com.

## Contributions

K.Y., N.D.-F., M.G.B., D.B., and J.T. contributed to the writing of the manuscript and generation of figures. K.Y., N.D.-F., Q.T., B.S., V.B., D.B., and J.T. contributed to the analysis of data. K.Y., N.D.-F., Q.T., S.B., J.A., B.S., D.L., N.P., and J.T. developed tools and methods that enabled the scientific discoveries herein. K.Y, N.D.-F., and J.T. contributed to the collection of the PhenoGenius dataset. All authors listed under PhenoGenius Consortium contributed to the generation of the primary data incorporated into the PhenoGenius resource. All authors reviewed the manuscript.

## Acknowledgments

We sincerely thank all patients, clinicians, biologists, bioinformaticians, and data scientists involved in this project. We are grateful to our scientific mentors for their helpful advice: Stanislas Lyonnet and Jean-Louis Mandel. Warm thanks to Jennifer Butt, who provided an extensive English language review of this manuscript. Special thanks are addressed to the Association Francophone de Génétique Clinique (https://af-gc.fr/) and to the physicians who performed phenotyping on the three clinical observations: Roseline Caumes, Mélanie Fradin, Anne-Marie Guerrot, Valentin Ruault, Godelieve Morel, Aurelia Jacquette, Gwenaël Le Guyader, Benjamin Dauriat, Geoffroy Delplancq, Bertrand Chesneau, Annick Toutain and Sarah Cluzel. This work has been partially supported by MIAI@Grenoble Alpes (ANR-19-P3IA-0003).

## PhenoGenius consortium

*Yannis Duffourd* ^*6*^, *Nicolas Chatron* ^*7*^, *Cedric Le Maréchal* ^*8*^, *Jean-François Taly* ^*9*^, *Wilfrid Carre* ^*10*^, *Claire Bardel* ^*11*^, *Frederic Tran Mau-Them* ^*6*^, *Marc Planes* ^*12*^, *Marie-Pierre Audrezet* ^*12*^, *Laure Raymond* ^*9*^, *Charles Coutton* ^*13*^, *Pierre Ray* ^*13*^, *Veronique Satre* ^*13*^, *Klaus Dietrich* ^*13*^, *Isabelle Marey* ^*13*^, *Françoise Devillard* ^*13*^, *Radu Harbuz* ^*13*^, *Florence Amblard* ^*13*^, *Pauline Le Tanno* ^*13*^, *Mouna Barat-Houari* ^*14*^, *Marjolaine Willems* ^*14*^, *Thomas Guignard* ^*14*^, *Sylvie Odent* ^*15*^, *Marie de Tayrac* ^*10*^, *Damien Sanlaville* ^*7*^, *Laurence Faivre* ^*6*^, *Laurent Mesnard* ^*16*^

^*6*^ Inserm UMR 1231 GAD, Genetics of Developmental disorders and Centre de Référence Maladies Rares Anomalies du Développement et syndromes malformatifs FHU TRANSLAD, Université de Bourgogne-Franche Comté, Dijon, France.

^*7*^ Department of Medical Genetics, Lyon University Hospital, 69677 Lyon, France; CNRS UMR 5292, INSERM U1028, CNRL, 69500 Lyon, France; Université Claude Bernard Lyon 1, GHE, 69100 Lyon, France.

^*8*^ Laboratoire de Génétique, UMR 1078 Génétique, Génomique fonctionnelle et Biotechnologies, Inserm, Université de Brest, EFS, CHU Brest, Brest, France.

^*9*^ Service de Génétique, Eurofins Biomnis, Lyon, France

^*10*^ Service de Génétique Moléculaire et Génomique, CHU Rennes, Rennes, France.

^*11*^ Univ Lyon, Université Lyon 1, CNRS, Laboratoire de Biométrie et Biologie Evolutive UMR5558, Villeurbanne, France. Service de Biostatistique-bioinformatique et plateforme NGS-CHU Lyon, Hospices Civils de Lyon, Lyon, France.

^*12*^ Laboratoire de Génétique, CHU Brest, Brest, France.

^*13*^ Genetic Epigenetic and Therapies of Infertility, Institute for Advanced Biosciences, Inserm U1209, CNRS UMR 5309, Université Grenoble Alpes, 38000, Grenoble, France. CHU de Grenoble, UM de Génétique Chromosomique, 38000, Grenoble, France.

^*14*^ Département de Génétique Médicale, Maladies Rares et Médecine Personnalisée, Univ Montpellier, CHU de Montpellier, CLAD ASOOR Montpellier, France.

^*15*^ Service de Génétique Clinique, Centre Référence “Déficiences Intellectuelles de causes rares” (CRDI), Centre de référence anomalies du développement CLAD-Ouest, CHU Rennes, 35203 Rennes, France; CNRS UMR 6290, Université de Rennes, 2 Avenue du Professeur Léon Bernard, 35043 Rennes, France.

^*16*^ Soins Intensifs Nephrologiques et Rein Aigu, Hopital Tenon, Assistance Publique - Hopitaux de Paris-Sorbonne Universite, Paris, France.

