## Supplementary Materials for "Learning phenotypic patterns in genetic diseases by symptom interaction modeling"

### Materials and Methods
### Supplementary Figures and Tables for

#### Learning phenotypic patterns in genetic diseases by symptom interaction modeling

*Kevin Yauy [1,2,*], Nicolas Duforet-Frebourg [2], Quentin Testard [1], Sacha Beaumeunier [2], Jerome Audoux [2], Benoit Simard [3], Dimitri Larue [2], Michael G.B. Blum [2], Virginie Bernard [4], David Genevieve [5], Denis Bertrand [2], PhenoGenius consortium, Nicolas Philippe [2], Julien Thevenon [1,4,*],*

1 Institute of Advanced Biosciences, Centre de recherche UGA, Inserm U 1209, CNRS UMR 5309, Grenoble, France.
2 SeqOne Genomics, Montpellier, France.
3 OuestWare, Nantes, France.
4 Centre de référence Anomalies du développement, CHU Grenoble-Alpes, Grenoble, France.
5 Department of Medical Genetics, Rare Disease, and Personalized Medicine, IRMB, University of Montpellier, National Institute of Health and Medical Research, Montpellier University Hospital Center, Montpellier, France.

**This PDF file includes:**

Materials and Methods
Figures S1 to S13
Tables S1 to S4

#### Materials and Methods

##### Clinical data collection

We collected anonymized clinical cases from four international cohorts leading to a total of 1,686 patients with a genetic diagnosis and their clinical description in HPO terms. This cohort is composed of 307 patients gathered from the PhenoGenius consortium from Centre hospitalier universitaire (CHU) Grenoble Alpes, CHU de Dijon, CHU de Montpellier, CHU de Rennes, CHU de Brest and Hospices Civils de Lyon, 140 patients from Seo *et al.* (*1*), 298 patients from Trujillano *et al.* (*2*), and 941 from Peng *et al.* (*3*). We also collected clinical descriptions in HPO format from 12 clinical geneticists from 12 different French hospitals (CHU Lille, CHU Montpellier, CHU Rennes, CHU Rouen, CHU La Réunion, CH Alençon, CHU Poitiers, CHU Limoges, CH Versailles, CHU Toulouse, and CHU Tours). Each physician extracted HPO terms from the same three clinical reports of patients with different diagnostic genes (*KMT2D, KMD6A,* and *C3*), one case each from three physicians in French hospitals (CHU Montpellier, CHU Grenoble Alpes, and APHP). Patients or legal guardians provided informed written consent for genetic analyses in a medical setting. Consent from the clinical geneticists was obtained through a survey that also collected their responses.

##### Database of medical literature

Databases were downloaded in May 2022. Human Phenotype Ontology was downloaded in OBO format from the Monarch Initiative website (https://hpo.jax.org/). Clinical databases in HPO format were downloaded from the Monarch Initiative website in the phenotype to genes format, EBI initiative DD2GP's (*4*) CSV files from https://www.ebi.ac.uk/gene2phenotype/, and Orphanet's XML data from https://www.orpha.net/. Free-text databases were downloaded through API requests for OMIM (https://www.omim.org/), NCBI's MedGen (*5*) (https://www.ncbi.nlm.nih.gov/medgen/) and NCBI's PubMed abstracts. For the PubMed abstracts, we used the list of all likely pathogenic and pathogenic variants from the ClinVar database (*6*) to select abstracts of potential interest through LitVar (*7*) API.

##### Text matching algorithm

###### Methods

We developed a methodology to extract symptoms-gene associations based on Elasticsearch® v5.6 from free-text data in HPO terms and NCBI gene ID format. We first processed these databases to associate free-text data with the corresponding gene in JSON format. An Elasticsearch query was performed to match every HPO available in each gene-free-text related data and provide a list of HPO-gene associations per database. We limited the number of gene-HPO associations created to the top 100 ranked associated genes for an HPO.

###### Mean distance in the ontology between exclusive terms

For each exclusive symptom-gene association from the MI database, we computed ontology distance for every exclusive symptom-gene association from text-mined OMIM in a common gene and kept the lowest distance. For each gene, we processed the mean distance in the ontology between exclusive terms of the MI database and our text-mined OMIM database. We performed the same experiment in the opposite direction, from exclusive association in text-mined OMIM database to MI database. To compare the distribution of mean distance against a random distribution, we computed ontology distance with a randomly selected HPO term instead of an exclusive term from the other database.

##### Knowledge data frame structure

A list of symptom-gene associations from each database was stored in a data frame containing 16,600 symptoms in columns and 5,235 genes in rows. Each cell includes the probability of symptom-gene association according to its overlap between databases (consensus score based on a mean, e.g. an association found in half of the databases received a score of 0.5). Databases structured in HPO format (MI, DDG2P, and Orphanet) were considered a unique resource, as DDG2P and Orphanet provided associations mostly overlapping with the MI database and with only 3,492 and 1,849 exclusive associations, respectively.

##### Node similarity

We transposed the symptoms-gene associations' data frame into a symptom-symptom association data frame based on symptoms association in the same genes. We injected all existing symptoms to symptoms relationships into a Neo4j® database v4.4.0. The similarity between all pairs of symptoms was processed using the node similarity algorithm (https://neo4j.com/docs/graph-data-science/current/algorithms/node-similarity/), and due to technical reasons (RAM limit due to number of combinations), for each symptom, we extracted symptoms with a similarity score > 0.4 and limited to a maximum of 1500 associated symptoms. A similar pair of symptoms was reported if the similarity score was higher than 0.8.

#### Collaborative filtering

##### Methods

Using sci-kit learn v.0.24.2 Non-Negative Matrix Factorization with Nonnegative Double Singular Value Decomposition initialization, we transposed the symptoms-gene associations data frame into a symptoms-groups of symptoms association data frame based on symptoms association in the same genes. This algorithm provides the numbers of groups requested, the weights of symptom-group associations, and the weight of gene-group associations. For interpretability and illustration, we filtered symptoms-group association keeping only the 10% highest l2-normalized weight of symptoms-group association (>0.04). In phenotype matching evaluation, we use the complete symptoms-group associations.

##### Identification of an optimal number of symptoms group

We applied a coherence score metric to processed groups of symptoms to select their optimal number. Using gensim v4.2 implementations of the coherence topic evaluation (https://radimrehurek.com/gensim/models/coherencemodel.html), we sought the range of group numbers with the highest coherence score and also the lowest coefficient of variation using five random state initialization. We looked for consistency of coherence among different random states to reproduce the same performance with additional data or updates. This is necessary for clinical implementation.

##### UMAP and clustering

Using sci-kit learn's v0.24.2 agglomerative clustering implementation (affinity="euclidean" and linkage="ward" parameters), we obtained hierarchical clusters. Data were normalized per clinical observations and also per group of symptoms. 75 clusters of clinical descriptions were retained to avoid single observation clusters. Dendrograms were obtained using Scipy v1.8. hierarchy module. UMAP visualization was performed using the umap v0.5.3 module, with the following parameters: neighbours = 3 & minimum distance = 0.9 for 390 groups of symptom dimension and neighbours = 2 & minimum distance = 0.9 for 16,600 symptoms dimension. We determined these parameters after observing dispersion and clustering of clinical descriptions according to a range of neighbors from (2 to 5) and minimum distance (0.1 to 0.99).

##### Graph visualization

Exploration plots were processed using python's plotnine v0.9 package. Graph visualization was obtained using software Gephi v0.9 (https://gephi.org/) with ForceAtlas2 (*8*) and Neo4j Bloom v2.3. Retina (https://ouestware.gitlab.io/retina/beta/) was used to provide users with a visual browser of graphs.

##### Phenotype matching

A phenotype match occurred if at least one symptom in the clinical description was related to the diagnostic gene in the database. We developed a phenotype matching system that matches clinical descriptions with lists of symptoms-gene associations available in the knowledge data frame structure. According to the combination of symptoms from clinical descriptions, the data frame was filtered to contain only selected symptoms or groups of symptoms columns. The sum of the consensus score per row or gene was processed, and genes ranked according to the sum score in descending order. In the case of equal scoring, we applied the worst rank to all equal genes. The evaluation of this phenotype matching using databases in HPO format and text-mined associations used only symptoms declared in the clinical description. The phenotype matching system based on symptoms similarity used declared symptoms and added a virtual symptom containing all highly similar symptoms sharing its weight. The method based on NMF's collaborative filtering projected symptoms into 390 groups of interacting symptoms dimension using a trained model. Each gene is also projected in the 390 groups dimension with different weights. The ranking is calculated based on the normalized Euclidean distance between the 390 group projection of each gene and the patient phenotypes. The gene with the highest distance gets the best ranking.

##### Comparisons of phenotype-driven gene prioritization systems

We benchmarked the performance of four different phenotype-driven gene prioritization algorithms: PhenoApt, Phen2Gene, CADA, and LIRICAL (*3*, *9*–*11*). As each tool provides a different maximum limit for the gene ranking list, we performed our diagnostic performance evaluation based on the top 100 cumulative causal gene ranks to be interpretable. PhenoApt (https://www.phenoapt.org/API) and Phen2Gene (https://phen2gene.wglab.org/api) evaluations were processed using the software's API in May 2022. CADA (https://github.com/Chengyao-Peng/CADA, unique release) and LIRICAL (https://github.com/TheJacksonLaboratory/LIRICAL, v.1.3.4) evaluations were processed using the desktop version via GitHub.

#### Statistics

Statistical metrics (Kolmogorov-Smirnov test, Spearman correlation coefficient, Fisher exact test) were obtained using Python's SciPy module v1.8. Benjamini Hochberg correction was obtained using the multipy package v0.16.

#### Methods references

9. Z. Chen, Y. Zheng, Y. Yang, Y. Huang, S. Zhao, H. Zhao, C. Yu, X. Dong, Y. Zhang, L. Wang, Z.

Zhao, S. Wang, Y. Yang, Y. Ming, J. Su, G. Qiu, Z. Wu, T. J. Zhang, N. Wu, PhenoApt leverages clinical expertise to prioritize candidate genes via machine learning. *Am. J. Hum. Genet.* **109**, 270–281 (2022).

10. M. Zhao, J. M. Havrilla, L. Fang, Y. Chen, J. Peng, C. Liu, C. Wu, M. Sarmady, P. Botas, J. Isla, G. J. Lyon, C. Weng, K. Wang, Phen2Gene: rapid phenotype-driven gene prioritization for rare diseases. *NAR Genom Bioinform*. **2**, lqaa032 (2020).

11. P. N. Robinson, V. Ravanmehr, J. O. B. Jacobsen, D. Danis, X. A. Zhang, L. C. Carmody, M. A. Gargano, C. L. Thaxton, UNC Biocuration Core, G. Karlebach, J. Reese, M. Holtgrewe, S. Köhler, J. A. McMurry, M. A. Haendel, D. Smedley, Interpretable Clinical Genomics with a Likelihood Ratio Paradigm. *Am. J. Hum. Genet.* **107**, 403–417 (2020).

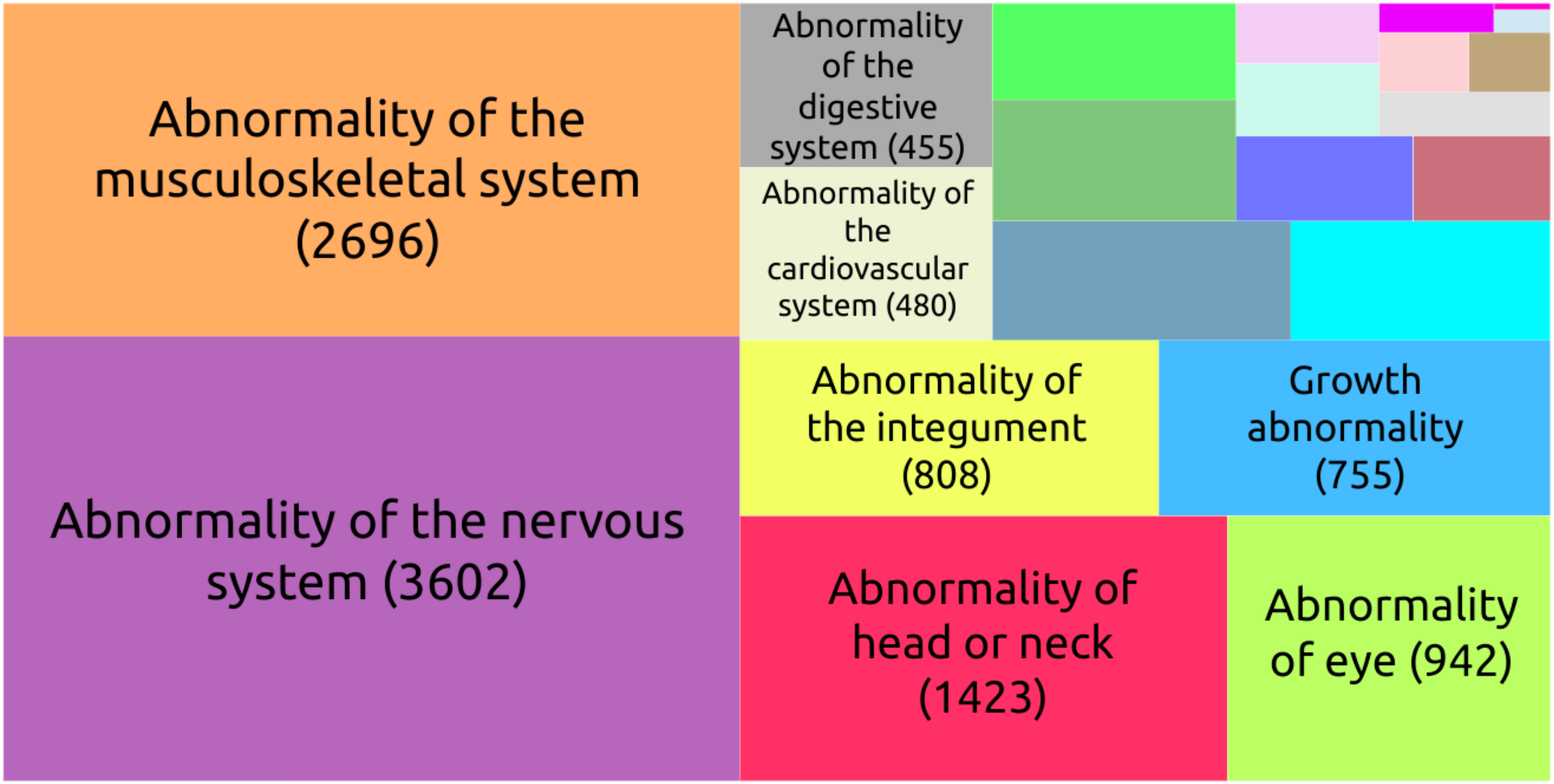


**Fig. S1. A. Treemap chart of the HPO terms in the cohort per main class ontology.**

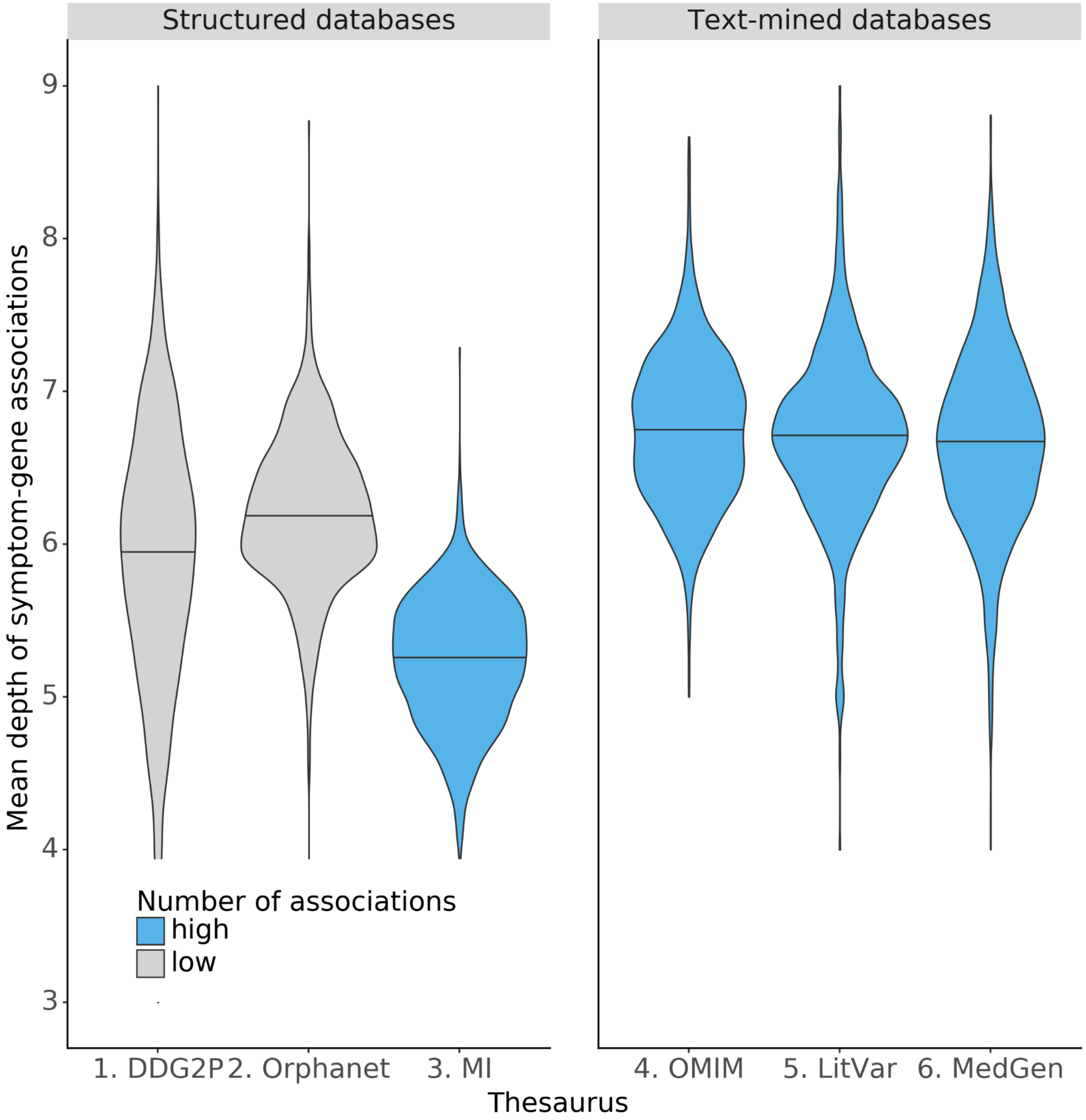


**Fig. S2. Violin plot of the mean depth of HPO terms from root ontology according to each database.**

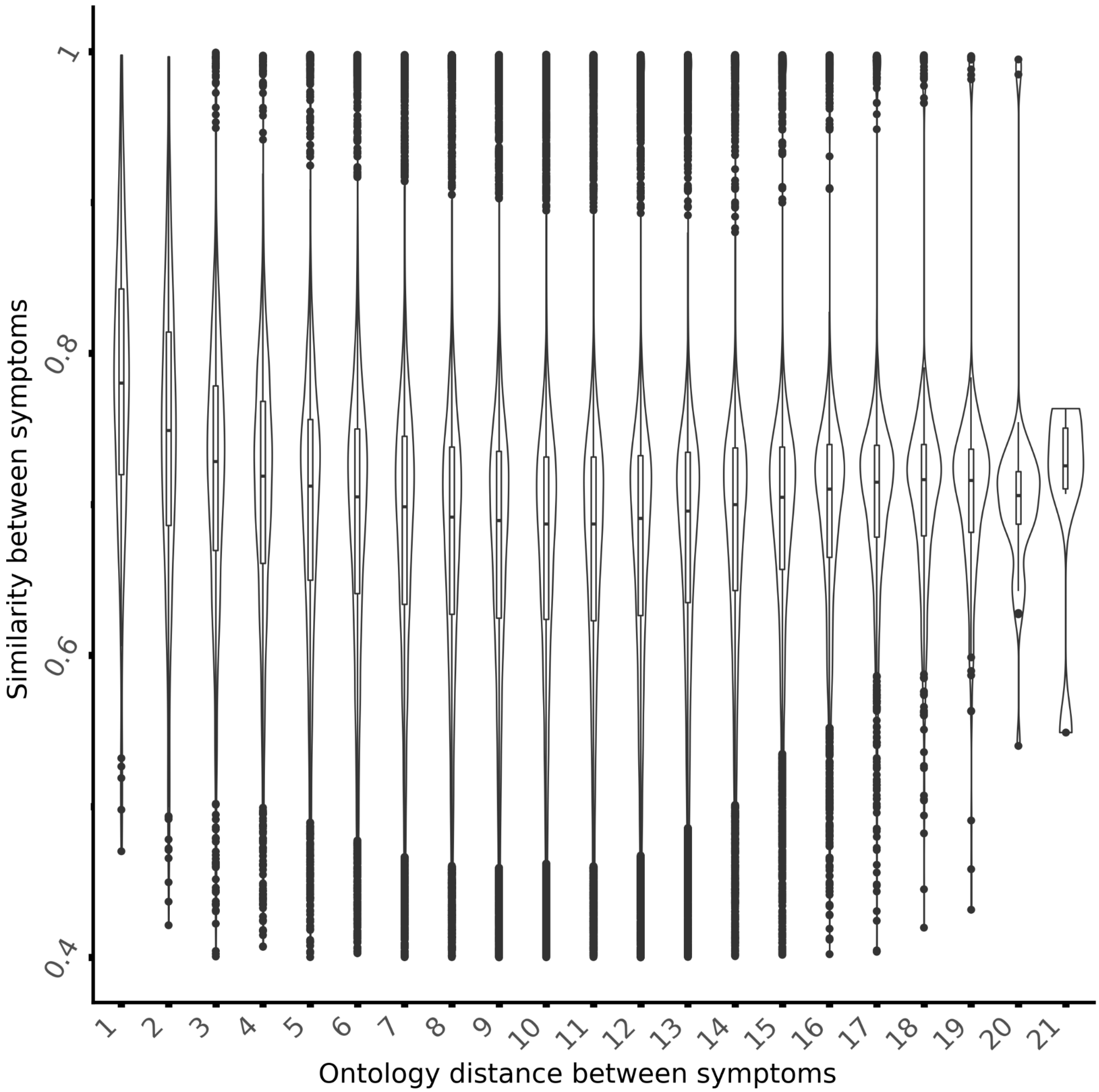


**Fig. S3. Violin plot of pair of symptoms similarity score according to the ontology distance.**

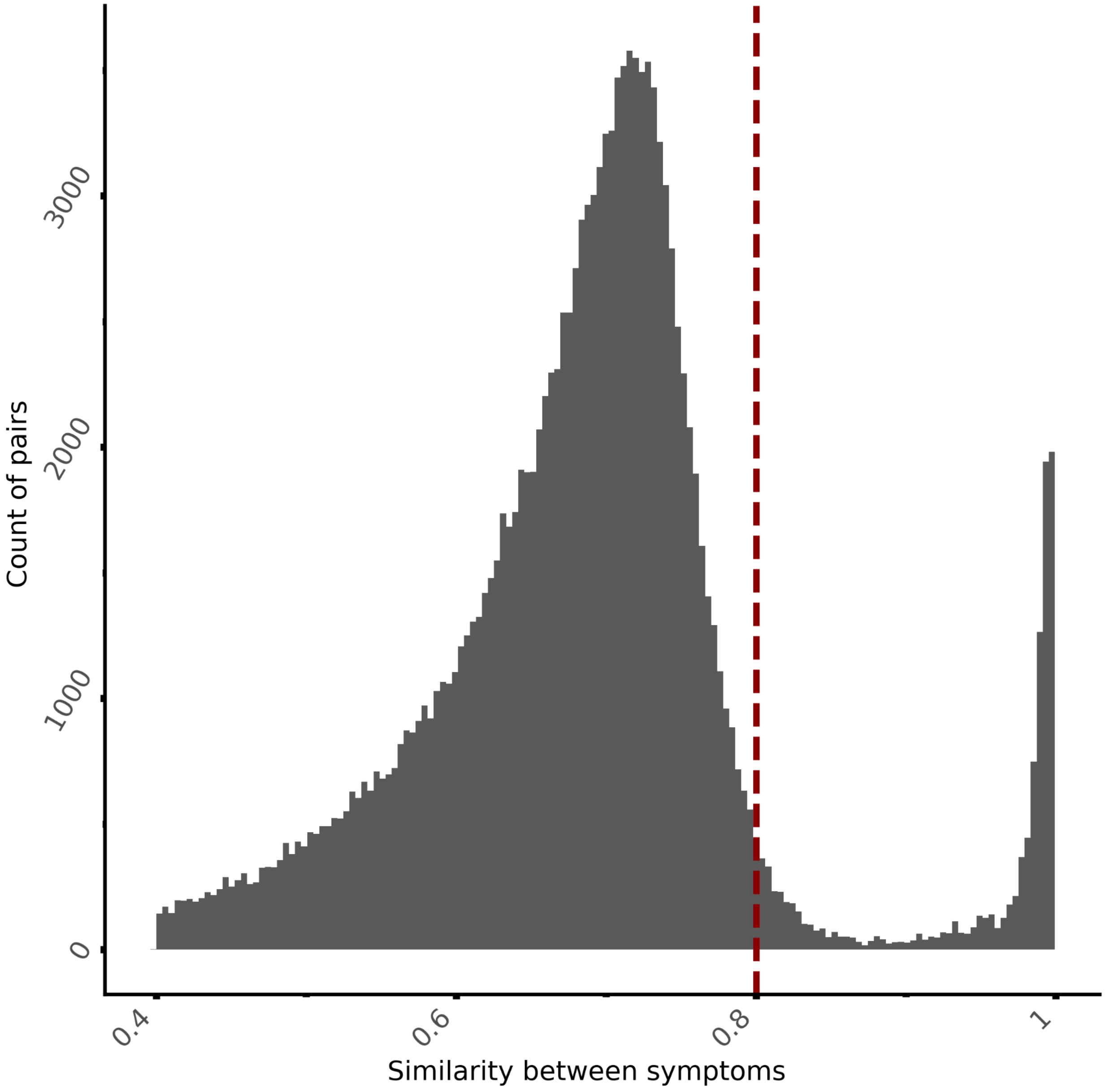


**Fig. S4. Distribution of 1% subsampling of similarity pair score.**

The dashed red line corresponds to the 80% similarity threshold and represents 10% of similarity pairs.

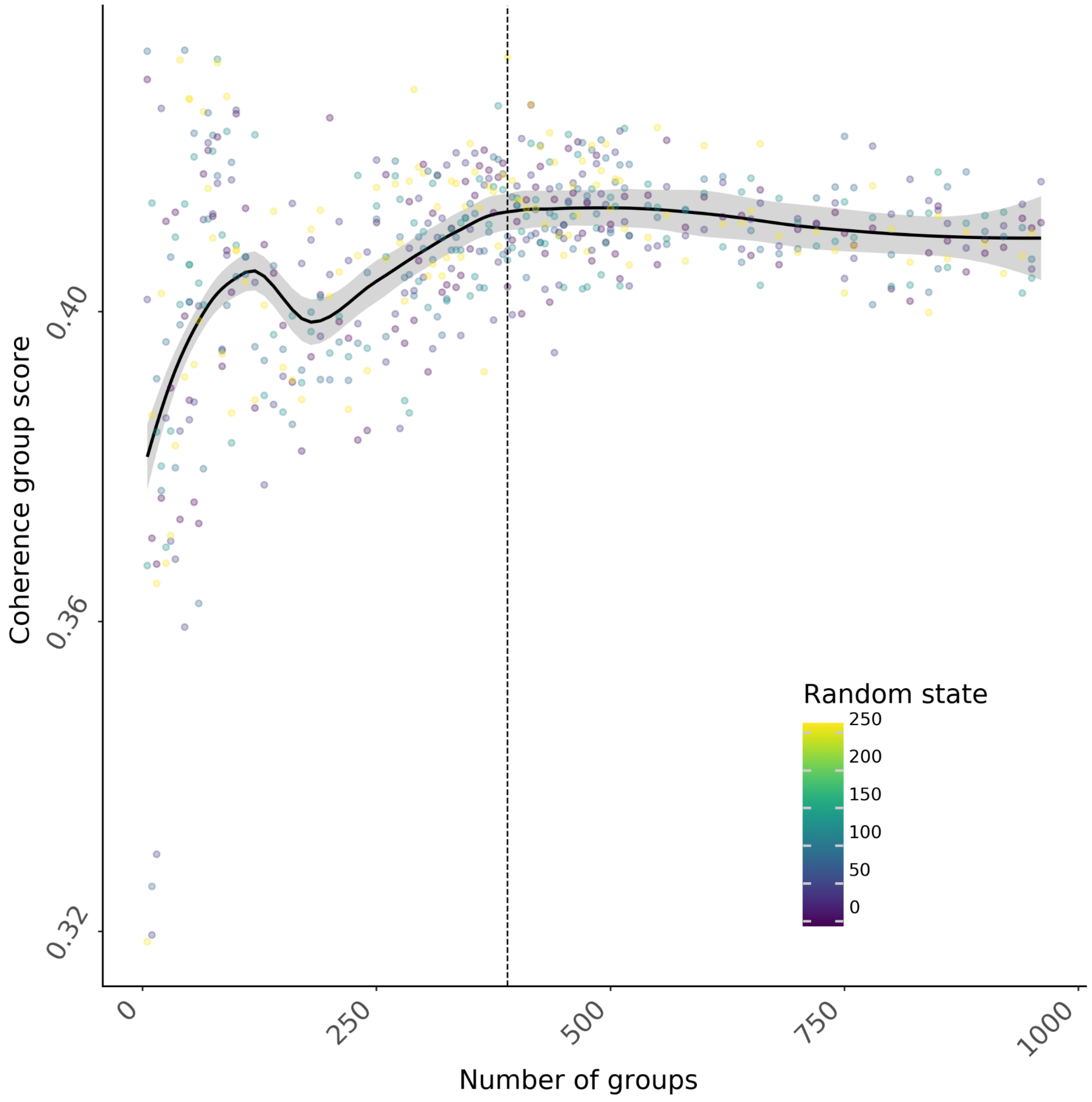


**Fig. S5. Topic coherence evaluation to determine the optimal number of groups.**
Colors represent iterative training experiments according to random state initialization.

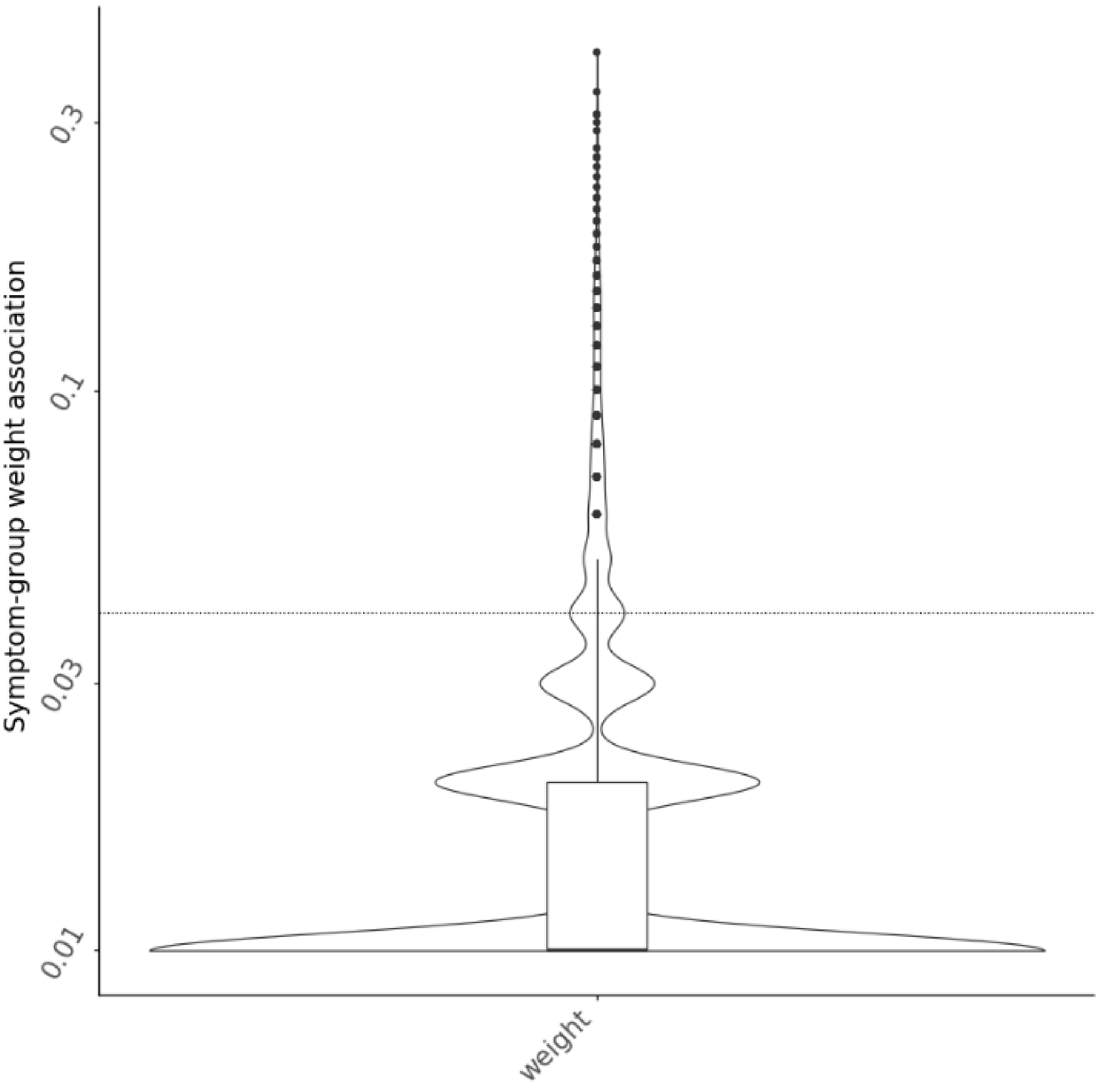


**Fig. S6. Violin plot of symptom-group weight association.**
Dashed horizontal line corresponds to the top 10% of symptom-group associations normalized weight threshold (>0.04).

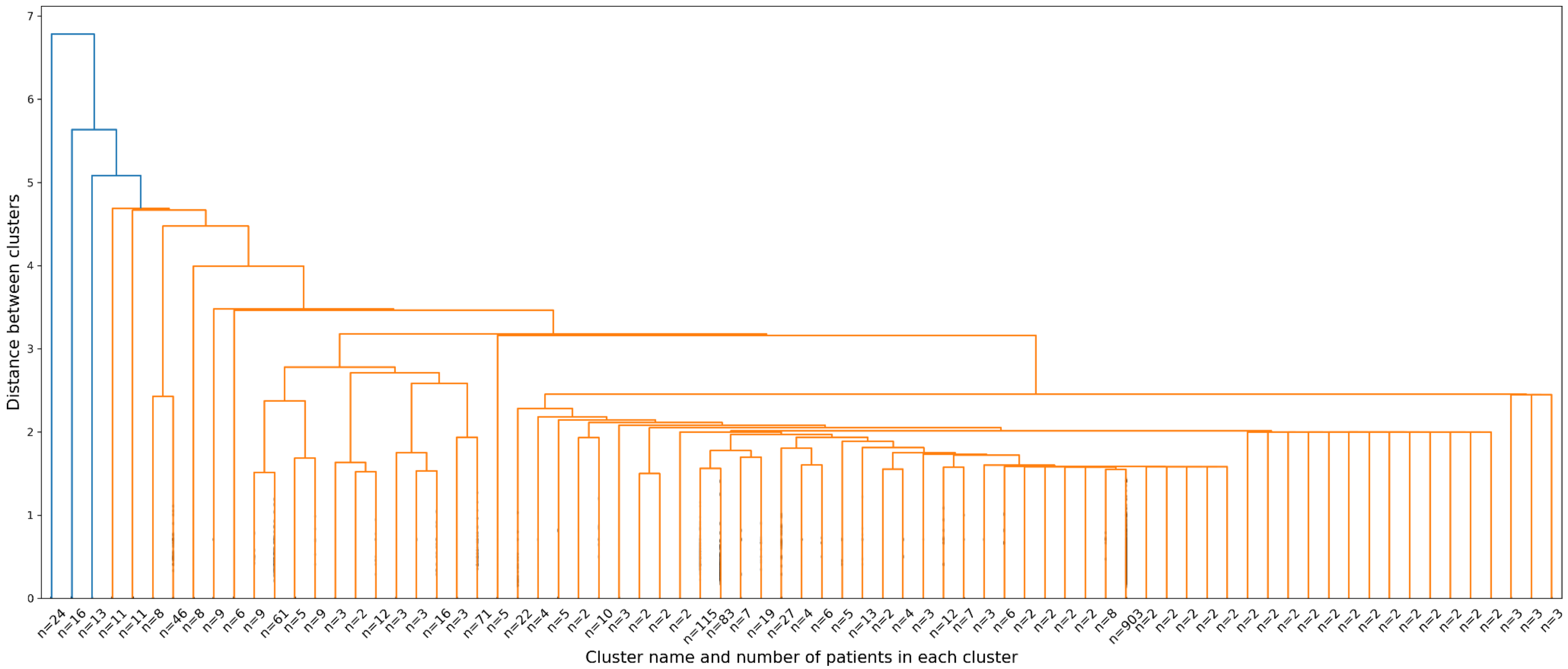


**Fig. S7. Dendrogram of hierarchical clusters of clinical observations obtained using agglomerative clustering on the retrospective cohort using the 16,600 symptoms from HPO.** The count of observations per cluster was reported (n). Colors represented branches of the hierarchy.

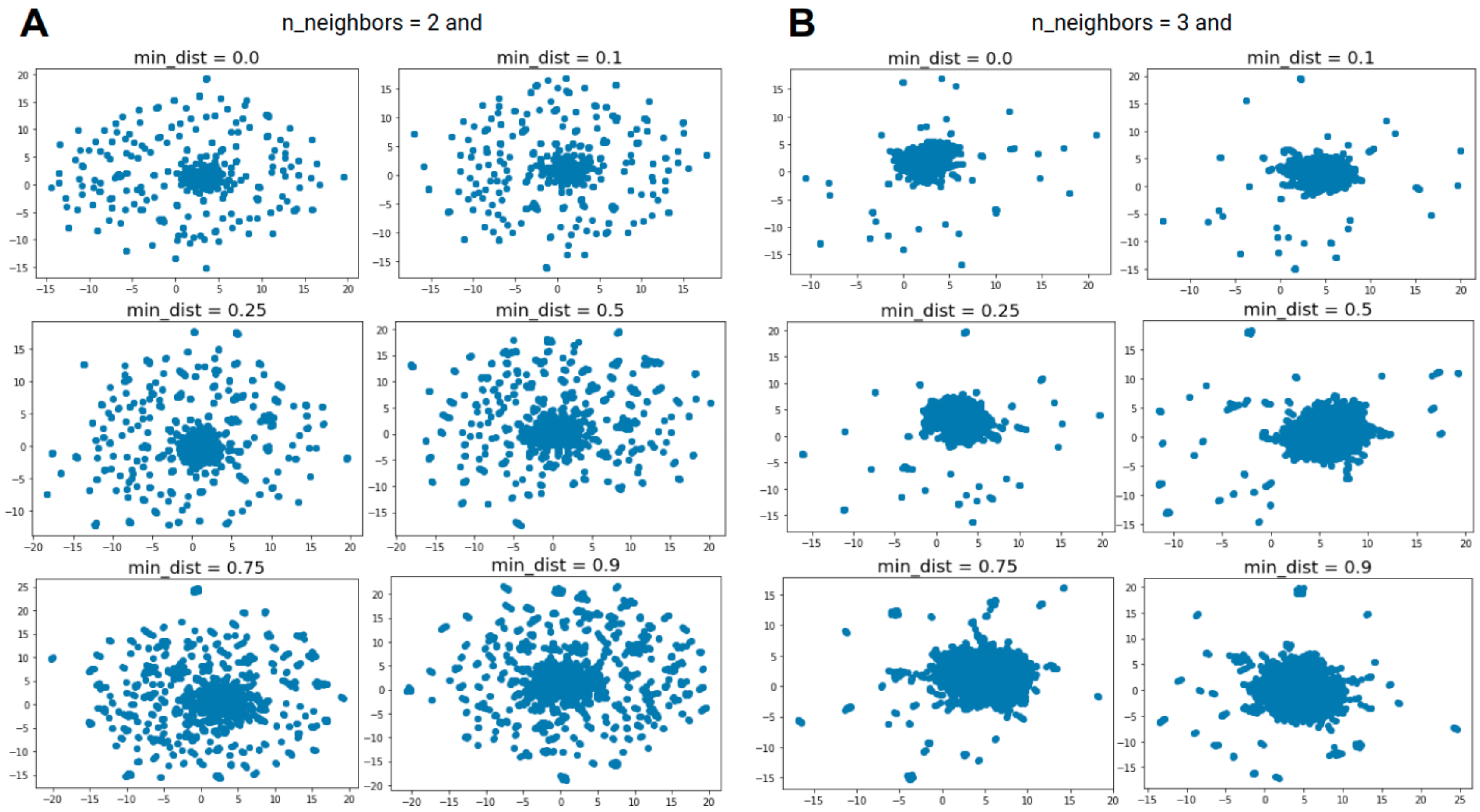


**Fig. S8. UMAP visualization of clinical description from the retrospective cohort of 1,686 cases using the 16,600 symptoms from HPO.**

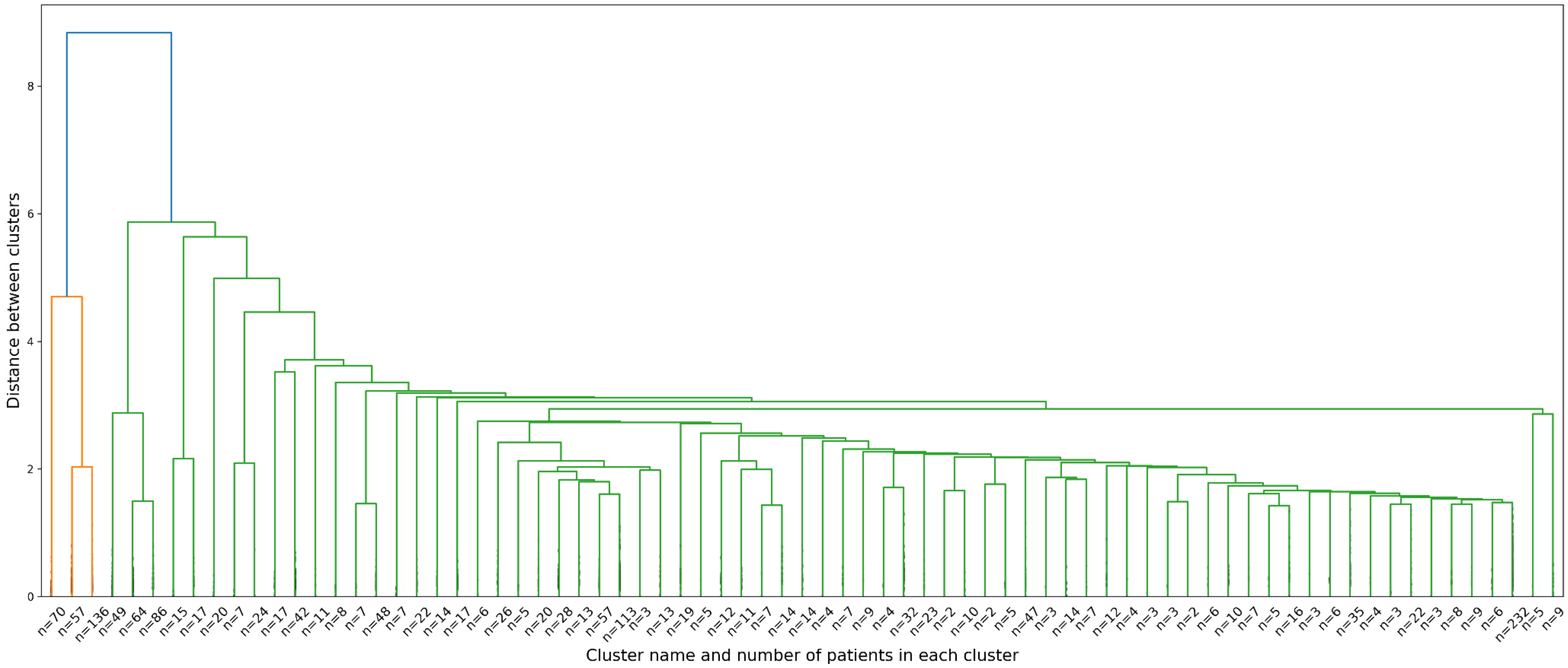


**Fig. S9. Dendrogram of hierarchical clusters of clinical observations obtained using agglomerative clustering on cohort projection in 390 groups of interacting symptoms dimension.** The count of observations per cluster was reported (n). Colors represent branches of the hierarchy.

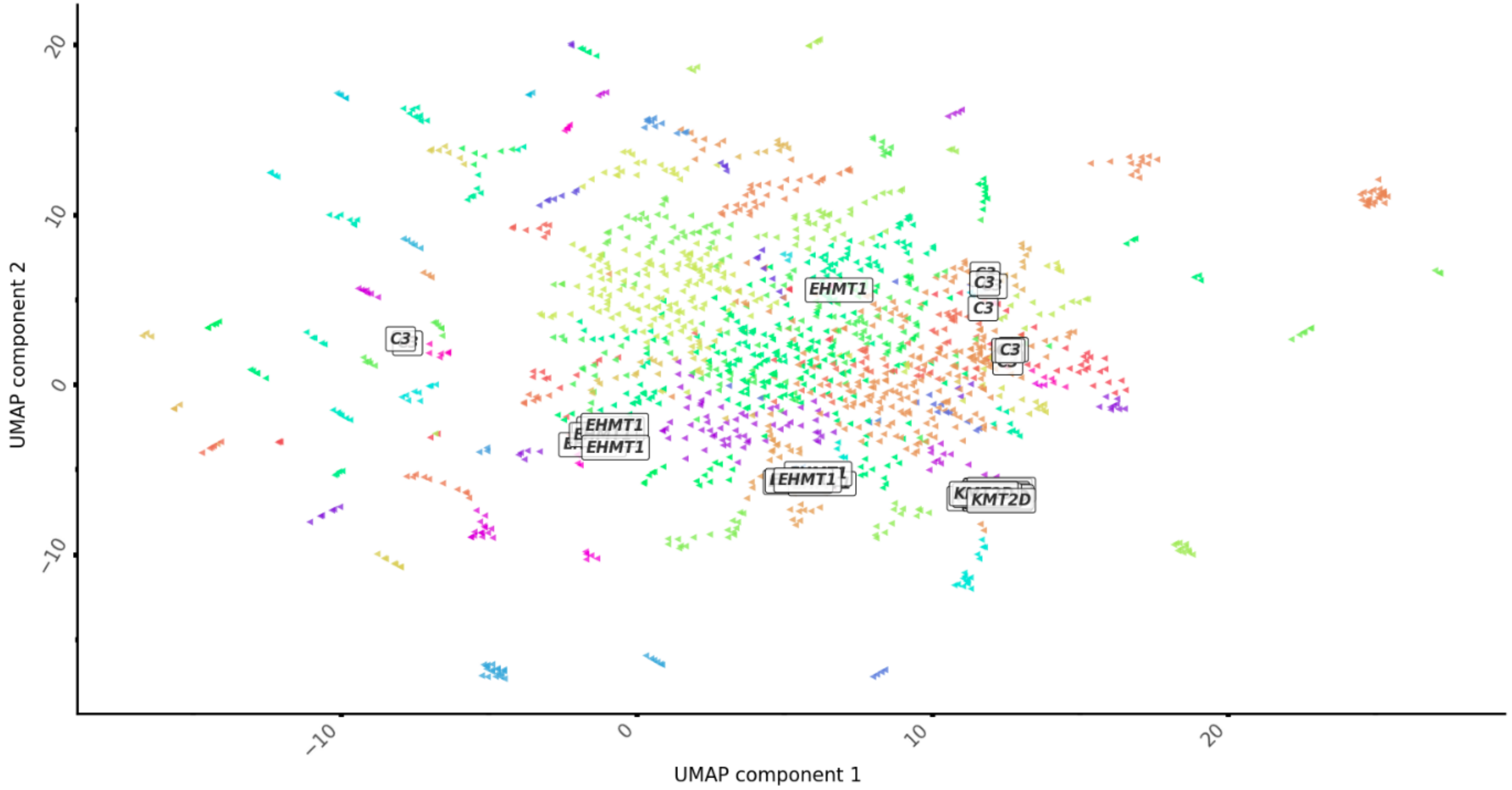


**Fig. S10. UMAP visualization of cohort's clinical descriptions projected using the 390 groups of interacting symptoms, colored and annotated by agglomerative cluster.** White boxes represent clinical reports description phenotyped by twelve physicians.

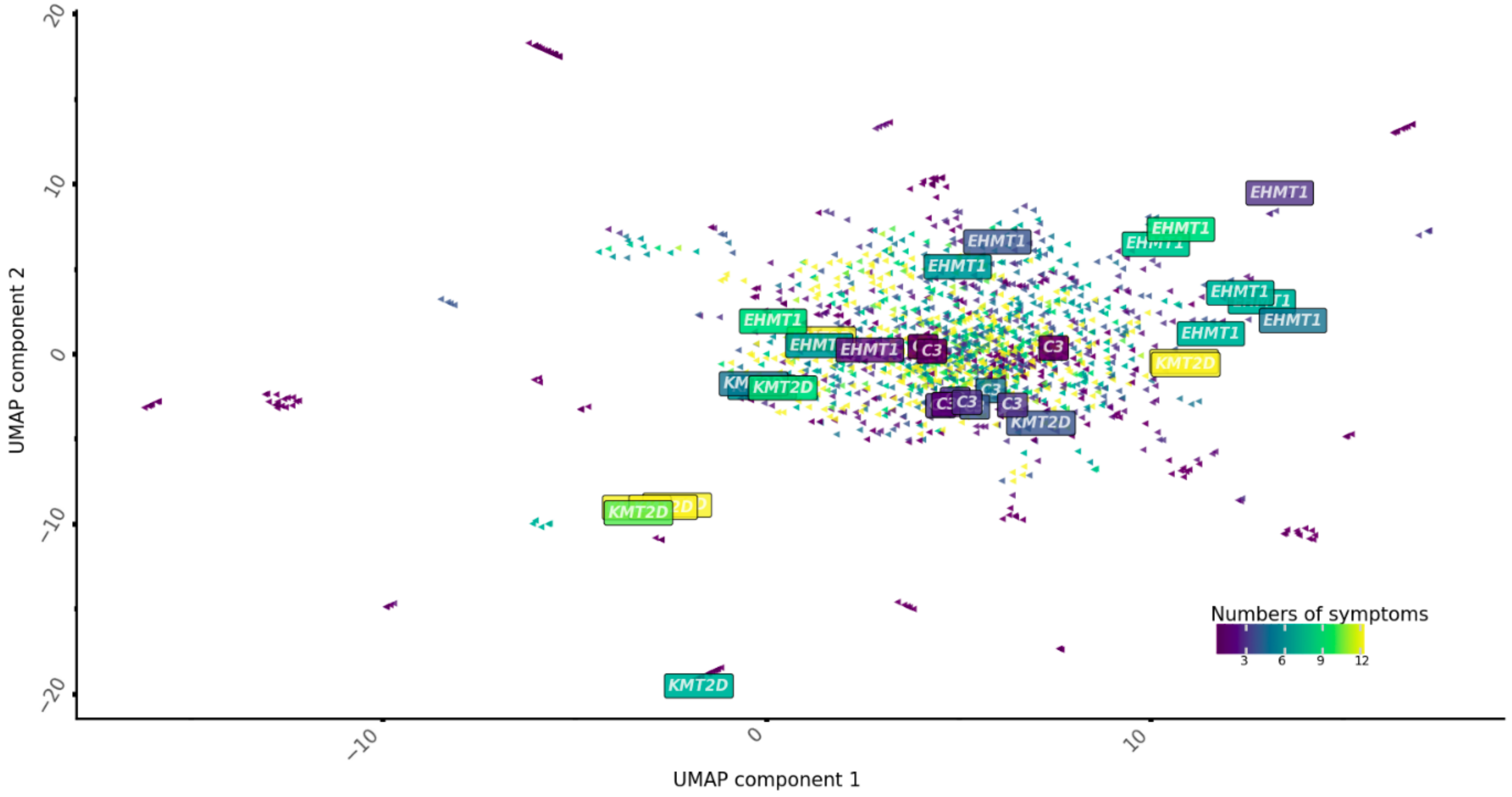


**Fig. S11. UMAP visualization of cohort's clinical descriptions projected using the 16,600 symptoms from HPO, colored by the number of symptoms.** Boxes represent clinical reports description phenotyped by twelve physicians.

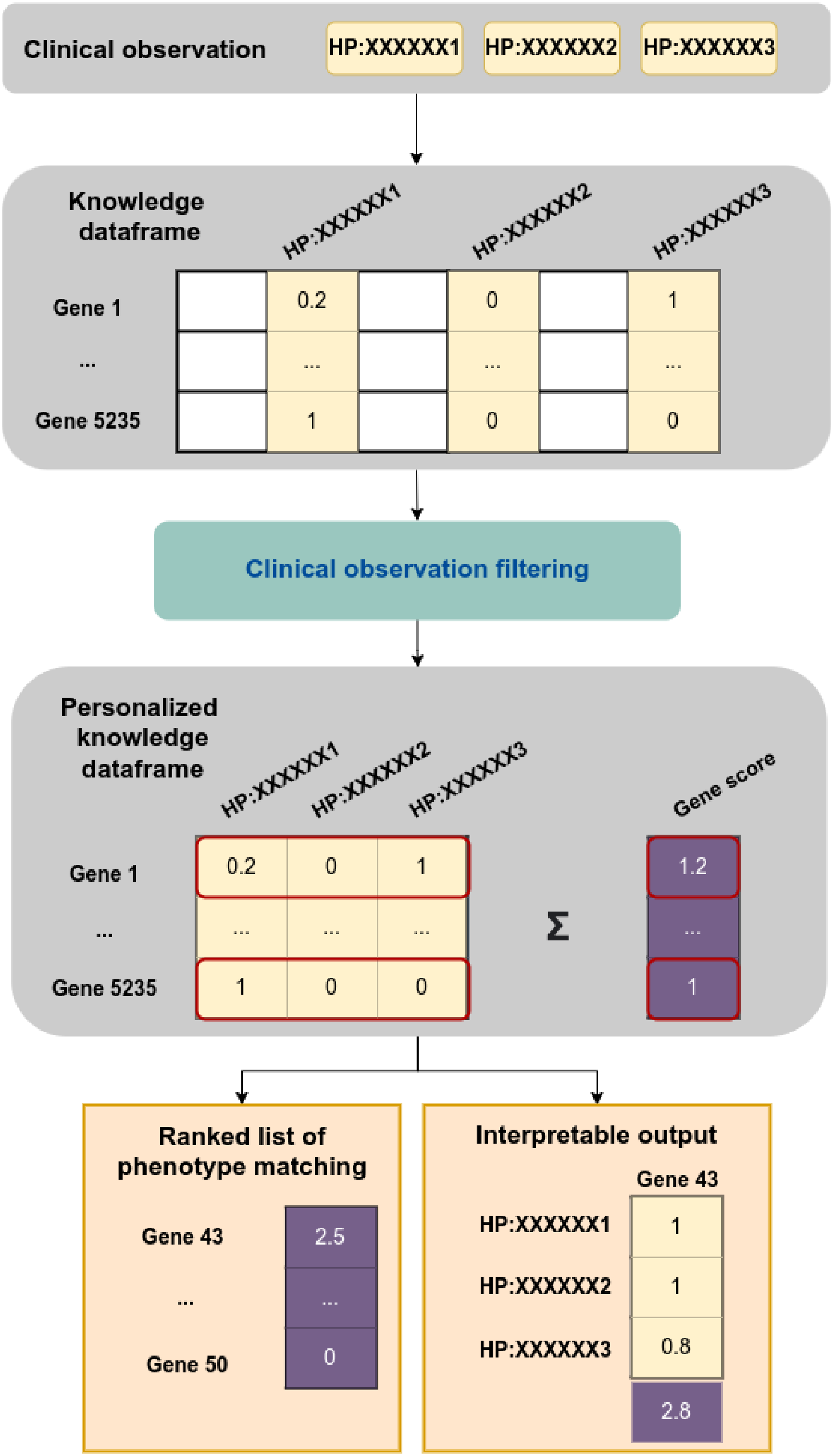


**Fig. S12. Illustration of phenotype matching and gene prioritization system.** According to symptoms of a clinical description in HPO format, the knowledge dataframe is filtered. A personalized and interpretable ranking of genes is provided according to the sum of associated symptom-gene associations available.

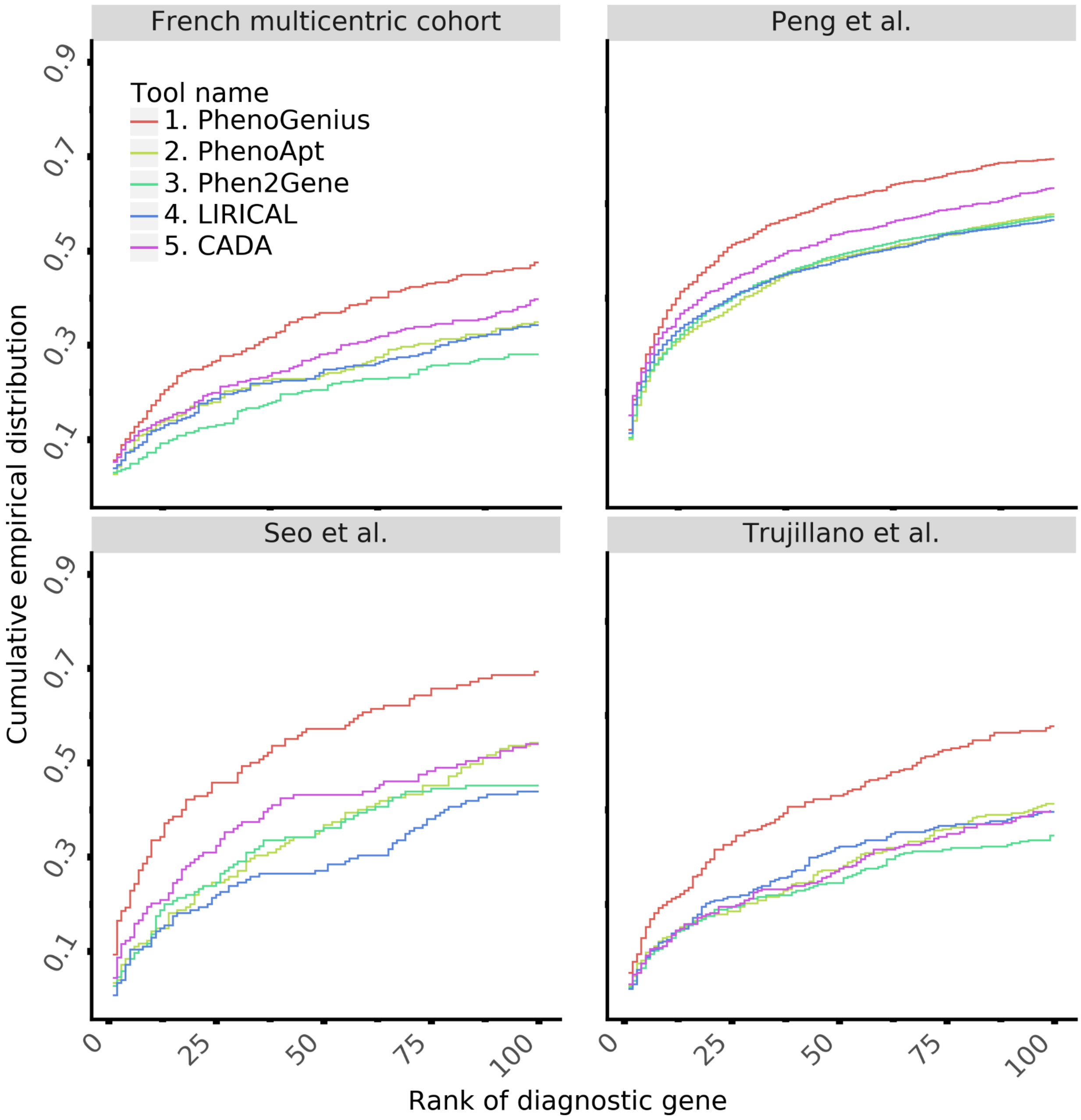


**Fig. S13. Benchmark of a selection of state-of-the-art phenotype-driven gene prioritization per sub-group cohort.**

The fraction of cases correctly diagnosed (y-axis) is plotted against a cumulative causal gene rank.

**Table S1. Clinical data collection of 1,686 patients in our international cohort.**

| ID | Description | Count of | | | |
|---|---|---|---|---|---|
| | | Patients | Genes | Terms Total | Unique |
| French multicenter cohort from PhenoGenius consortium | Gathered from CHU Grenoble Alpes, CHU de Dijon, CHU de Montpellier, CHU de Brest, and Hospices Civils de Lyon | 307 | 220 | 3243 | 989 |
| Seo *et al.* (*1*) | Unselected series of consecutive patients, clinically suspected of carrying a genetic disorder, from non-consanguineous families, who presented at the Medical Genetics Center, Asan Medical Center, Seoul, South Korea, from April 2018 to August 2019. | 140 | 120 | 1135 | 347 |
| Trujillano *et al. (2)* | Consecutive, unrelated patients referred by physicians from 54 countries on different continents have been included in this study. All patients with suspected Mendelian disorders were referred for diagnostic exome sequencing between January 2014 and January 2016. | 298 | 241 | 2411 | 789 |
| Peng *et al.* (*3*) | Collection of 435 descriptions from German hospitals and 506 ClinVar submissions. | 941 | 528 | 6439 | 1814 |

**Table S2. Top ten recurring genes in the four groups in our cohort.**

| Gene name | Count (n=1,686) | Percentage |
|---|---|---|
| *ABCC6* | 22 | 1.29 |
| *ANKRD11* | 21 | 1.23 |
| *ARID1B* | 20 | 1.18 |
| *NSD1* | 18 | 1.06 |
| *BLM* | 16 | 0.94 |
| *FBN1* | 15 | 0.88 |
| *MECP2* | 15 | 0.88 |
| *NF1* | 15 | 0.88 |
| *PTPN11* | 14 | 0.82 |
| *PKD1* | 13 | 0.76 |

**Table S3. Top ten recurring HPO terms in the four groups in our cohort.**

| HPO | Description | Count (n=13,228) | Percentage |
|---|---|---|---|
| HP:0001263 | Global developmental delay | 373 | 2.82 |
| HP:0000750 | Delayed speech and language development | 241 | 1.82 |
| HP:0001249 | Intellectual disability | 231 | 1.75 |
| HP:0000252 | Microcephaly | 209 | 1.58 |
| HP:0001250 | Seizure | 205 | 1.55 |
| HP:0001252 | Hypotonia | 170 | 1.29 |
| HP:0004322 | Short stature | 168 | 1.27 |
| HP:0001270 | Motor delay | 126 | 0.95 |
| HP:0001622 | Premature birth | 108 | 0.82 |
| HP:0000486 | Strabismus | 102 | 0.77 |

**Table S4. Description of clinical geneticist profiles in the prospective phenotyping experiment.**

| ID | Profile | Self-estimated expertise in phenotyping using HPO format (from 1 to 10) |
|---|---|---|
| 1 | Clinician | 7 |
| 2 | Clinician | 1 |
| 3 | Clinician | 5 |
| 4 | Resident in medical genetics | 9 |
| 5 | Clinician | 5 |
| 6 | Clinician | 1 |
| 7 | Clinician and laboratory specialist | 1 |
| 8 | Clinician | 6 |
| 9 | Clinician and laboratory specialist | 5 |
| 10 | Resident in medical genetics | 6 |
| 11 | Clinician | 3 |
| 12 | Resident in medical genetics | 1 |
